# Plasma and follicular fluid concentrations of carotenoids, tocopherols and retinol in a French population of women undergoing in vitro fertilization: a monocentric non-interventional study

**DOI:** 10.64898/2026.08.28.26360803

**Authors:** Astelle Ndiaye, Anne C.M. Thiébaut, Patrick Borel, Charlotte Sabran, Sébastien Elis, Fabrice Guerif, Virginie Maillard

**Author notes:** Corresponding authors: (VM), (ACMT).

## Abstract

The distribution of fat-soluble compounds (including antioxidants) in follicular fluid (FF) remains sparsely documented in relation to in vitro fertilization (IVF) outcomes and existing studies have reported diverging associations. This study aimed to describe plasma and FF concentrations of fat-soluble micronutrients in women undergoing IVF and to analyze their adjusted associations with ovarian function, embryo development and pregnancy outcomes. In 2021-2022, plasma and FF samples were collected from 82 women (first IVF cycle) at oocyte puncture, along with lifestyle data covering the three preceding months. Eleven compounds (two tocopherols, three xanthophylls, five carotenes and retinol) were quantified.

All compounds were detected in both compartments (lowest in FF) except phytoene, undetectable in FF. Plasma and FF α-tocopherol concentrations were positively associated with plasma estradiol levels before oocyte puncture (both p<0.01) while FF α-carotene and lycopene were inversely associated with plasma progesterone concentrations (p=0.01 and 0.02, respectively). Plasma phytofluene and phytoene were positively associated with mature oocyte rate (p=0.03 and p=0.01, respectively), while FF retinol was negatively associated (p=0.03). Carotenes, tocopherols and retinol were inversely associated with later IVF outcomes: fertilization rate (p<0.001 for plasma γ-tocopherol, 0.02 for FF retinol), top-quality embryo (p=0.02 for plasma phytofluene), biochemical pregnancy at day 7 post-embryo transfer (p=0.05 for plasma α-tocopherol, 0.02 for plasma α-carotene), clinical pregnancy (p=0.03 for plasma α-tocopherol, 0.01 for plasma phytoene) and live birth (p=0.04 for plasma α-tocopherol, 0.02 for plasma phytoene). Plasma and FF γ-tocopherol were positively associated with embryo fragmentation (both p<0.05). Finally, among xanthophylls, only plasma β-cryptoxanthin was positively associated with plasma progesterone concentrations (p=0.02).

Our findings of heterogeneous associations between tocopherols, carotenes, retinol and IVF outcomes across the stages of IVF suggest a beneficial effect limited to early outcomes and support a complex and context-dependent role of these compounds in female reproduction.

## INTRODUCTION

Human reproduction is declining worldwide, with more than half of countries now below replacement level [1]. Infertility, defined as the failure to achieve a clinical pregnancy after 12 months of regular unprotected intercourse, affects 8–12% of reproductive-aged couples globally [2]. In this context, identifying modifiable determinants of reproductive potential has become a major public health concern. Female fertility is influenced by a complex interplay of genetics, hormonal, environmental and lifestyle factors [3], among which age [4,5], obesity [6] and smoking [7] are well-established contributors, notably through their effects on ovarian reserve and oxidative stress.

Nutritional factors have been proposed as potential modulators of reproductive function [8,9]. In particular, vitamin A, a fat-soluble micronutrient, has been involved in reproductive processes. It is obtained either from provitamin A carotenoids such as β-carotene, α-carotene and β-cryptoxanthin and from preformed vitamin A (retinol and retinyl esters), found in diet, especially in animal products, fortified foods and supplements (for review, [10]). Retinol, through its active metabolite, retinoic acid, plays a critical role in meiosis initiation, ovarian function and early embryonic development (for review, [11,12]). Carotenoids and tocopherols (other fat-soluble compounds), which are exclusively obtained through the diet [13,14], have also been involved in female reproduction. Experimental studies have shown that vitamin E and several carotenoids, including β-carotene, lycopene, zeaxanthin and lutein, support oocyte maturation, follicular development and early embryo development in mammalian models, both in vitro [15–22] and in vivo [23,24]. The main biological mechanisms proposed for these molecules are their provitamin A activity for some of them or their antioxidant properties.

By modulating oxidative balance, antioxidant compounds such as carotenoids and tocopherols may influence the levels of reactive oxygen species (ROS) within the ovarian microenvironment, thereby affecting key processes involved in female reproduction [25]. Indeed, ROS production plays a dual role in female reproduction. At physiological levels, ROS are involved in key processes such as oocyte maturation, corpus luteum formation, and luteolysis (for review, [26–28]). However, excessive ROS accumulation may disrupt the follicular microenvironment. In follicular fluid (FF), oxidative imbalance has been associated with impaired fertilization, embryo development, and implantation, ultimately leading to poorer assisted reproductive technology (ART) outcomes (for review, [28,29]). Importantly, the effects of ROS appear to be stage-dependent, highlighting the need to better characterize their role across reproduction phases and the roles of molecules, including antioxidants, able to control their production.

In humans, several studies have investigated associations between fat-soluble compounds (vitamin E, carotenoids and vitamin A) and fertility through dietary assessment. While some studies have reported positive associations between dietary intakes and reproductive outcomes, including a lower infertility probability (vitamins A and E, lutein/zeaxanthin, lycopene, [30,31]), a normal cleaved embryo rate (retinol, [32]), a shorter time to pregnancy (β-carotene, [33]) and higher antral follicle count (AFC: lycopene, [34]), others have found no significant associations for some compounds [32,35,36] and some have even reported inverse relationships with certain reproductive outcomes, including AFC (retinol, [34]) and live birth rate (β-carotene, [36]). These discrepancies may reflect inherent limitations of dietary data, which do not account for inter-individual differences in absorption, metabolism or tissue distribution, and may not accurately reflect bioavailable concentrations in plasma or in the ovarian microenvironment. This underscores the need for studies based on direct biomarker measurements.

To date, few studies have investigated the relationship between fat-soluble micronutrients from plasma or serum and reproductive outcomes. Some have found no significant associations with certain compounds and oocyte maturation (vitamin E, [37]), fertilization success (retinol and α-tocopherol, [38]) and pregnancy status (retinol, lutein, lycopene, [39,40]), α-tocopherol, α- and β-carotenes, β-cryptoxanthin, [39]), while another study reported positive associations with numbers of retrieved and mature oocytes (α-tocopherol, [40]) and pregnancy status (α-tocopherol, β-carotene, [40]).

Furthermore, since the composition of FF is closely related to oocyte competence [41], it is relevant to investigate the composition of this fluid with regard to these fat-soluble compounds. However, the contribution of these micronutrients within this environment remains poorly documented. To date, only a limited number of studies have measured FF levels of carotenoids, vitamins E and vitamin A in women undergoing ART. While some studies have reported negative associations between FF concentration of certain compounds and reproductive outcomes, such as pregnancy status (retinol, [40]) and embryo fragmentation (α-tocopherol, β-cryptoxanthin, [42]), others have observed positive relationships with numbers of mature oocytes and blastocysts, embryo quality (vitamin E, [43]), and pregnancy status (α-tocopherol, β-carotene, [40]) and several studies have found no significant relationships for all these compounds [39] or specifically for vitamins A and E [38], lutein/zeaxanthin [40,42], lycopene [40,42] and β-carotene [42]. None of the biomarker studies have included phytoene and phytofluene, colorless carotenoid precursors with notable antioxidant capacity, similar to that of α-tocopherol [44]. Furthermore, these studies have examined only a limited number of IVF outcomes simultaneously, even though micronutrients may exert stage-dependent effects on reproductive function.

In this context, the present study aimed to comprehensively characterize carotenoids, vitamin E and vitamin A profiles in both plasma and FF of women undergoing ART and to investigate their associations with ovarian response, oocyte competence, embryo development, and clinical IVF outcomes. To this end, 11 compounds, including α- and γ-tocopherols, lutein, zeaxanthin, β-cryptoxanthin, α- and β-carotene, lycopene, retinol, phytoene and phytofluene, were quantified in plasma and FF samples from 82 women undergoing IVF.

## METHODS

### Study design

This non-interventional monocentric study (called FERTENOX study) included 82 women undergoing ART between December 9, 2021 and July 11, 2022 at the Department of Reproductive Medicine and Biology of Regional University Hospital Centre (CHRU) of Tours, France. This study was approved by the ethics committee “Comité de Protection des Personnes”, Île-de-France II (ID RCB: 2020-A03241-38). All participants signed a written non-objection form, in accordance with the ethics committee, in view of the research category which did not require informed consent form (non-interventional research involving human participants [RIPH3] as defined in article L.1121-1 of the French “Code de la Santé Publique”). The study was prospectively registered on ClinicalTrials.gov under identifier NCT04866329. Women aged 18 to 42 years undergoing their first oocyte puncture for IVF were eligible. Exclusion criteria included opposition to data processing, oocyte or sperm donation, IntraCytoplasmic Sperm Injection (ICSI) with testicular sperm extraction or fertility preservation sample. Participants were informed about the study during the routine monthly IVF information session held approximately three months prior to oocyte retrieval. Eligibility was confirmed during pre-IVF bacteriological screening (1–2 months before oocyte puncture); at this time women completed a detailed lifestyle questionnaire. A chronological overview of the study design is provided in **S1 Fig**.

### Ovarian stimulation and IVF procedures

All participants underwent the standard ovarian stimulation protocol used at the CHRU of Tours. First, the desensitization of the hypothalamo-pituitary axis was achieved using either a gonadotropin-releasing hormone (GnRH) agonist (Decapeptyl®, n=6) or a GnRH antagonist (Orgalutran®, n=76), depending on the woman’s risk of ovarian hyperstimulation. Then, the ovarian follicle development was induced with the administration of follicle-stimulating hormone (FSH) alone (n=49) or in combination with luteinizing hormone (LH) (n=33). Finally, once follicles reached the appropriate size, ovulation was induced with human chorionic gonadotropin (hCG, Ovitrel®), carefully timed to trigger ovulation within 36 hours, synchronizing with the oocyte puncture process. A combined estrogen-progestin pre-treatment (Minidril® or Leeloo®) was administered in 36 women to optimize follicular synchronization prior to stimulation.

Oocyte puncture (day 0, **S1 Fig**) was performed under local or general anesthesia, while fasting. Punctured oocytes were processed according to standard IVF laboratory procedures. Fertilization was performed on the day of oocyte puncture, either by conventional IVF or by ICSI, depending on infertility etiology. Two days after oocyte puncture (day 2), embryos were cultured until day 5. The transfer of one or two fresh blastocyst(s) was performed on day 5 when feasible (**S1 Fig**). In specific clinical situations (e.g., ovarian hyperstimulation syndrome risk, hemoperitoneum, COVID-19 positive test, or technical difficulties during transfer), embryo transfer was postponed and performed using cryopreserved day-5 blastocysts. Following each first embryo transfer, women were monitored to assess pregnancy outcomes.

### Assessment of oocyte quality, embryo development and pregnancy success outcomes

Biological and clinical outcomes were assessed for each woman at key stages of the IVF process. The mature oocyte rate was defined as the number of intact metaphase II oocytes (extrusion of the first polar body) divided by the total number of collected oocytes. For ICSI cycles, this assessment was performed on day 0 (day of oocyte puncture), and for conventional IVF on day 1. The fertilization rate was defined as the proportion of oocytes presenting two pronuclei on day 1 relative to the number of mature oocytes. On day 2, embryo quality was assessed. The top-quality embryo rate was defined as the proportion of embryos with four regular blastomeres without multinucleation and with less than 20% cytoplasmic fragmentation. The fragmentation rate, on day 2, corresponded to the proportion of embryos exhibiting fragmentation among the number of embryos. The blastocyst rate was calculated as the number of day-5 blastocysts divided by the number of day-2 embryos. Additionally, the top-quality blastocyst rate on day 5 was defined as the proportion of top-quality blastocysts (full blastocysts with well-organized trophectoderm and the inner cell mass both graded [A or B] according to the Gardner’s grading guide [45]), relative to the number of blastocysts.

Early pregnancy was considered successful when blood β-hCG concentration 7 days post embryo transfer (day 7 Post Embryo Transfer [PET]) exceeded 5 IU/L, followed by confirmation (doubling of β-hCG concentration between 7- and 9-days PET). Biochemical pregnancy was confirmed on day 14 PET when β-hCG concentration exceeded 1000 IU/L. Clinical pregnancy, reflecting successful implantation, was defined by the presence of at least one fetus with discernible heartbeat, confirmed by ultrasonography at 8 weeks of amenorrhea. Live births were recorded for all completed pregnancies.

### Biological fluid collection

For each woman, plasma samples were collected the day before oocyte puncture during the routine blood test conducted at the end of ovarian stimulation monitoring. FF was collected the day of oocyte puncture (**S1 Fig**). To protect samples from oxidation, tubes were closed under nitrogen gas and immediately snap-frozen into liquid nitrogen, then stored at -80°C until analysis.

### Questionnaire and lifestyle data

Women completed a standardized questionnaire assessing lifestyle factors potentially influencing their redox status, including sociodemographic characteristics, smoking status, physical activity, dietary vitamin and mineral supplement use, and frequency of fruit and vegetable consumption over the previous three months. Given that fruits, vegetables and dietary supplements are major contributors of carotenoid and vitamin intakes, these data were collected for their potential relationship with measured plasma and FF concentrations of these molecules. The questionnaire also included self-reported height and weight, allowing the calculation of body mass index (BMI) (kg/m²).

### Analyses of carotenoids, tocopherols and retinol in plasma and follicular fluid

Extractions were performed on 250 µL of plasma and 500 µL of FF at room temperature (25°C ± 3°C) and under dim light to minimize light-induced isomerization of the compounds of interest, following the method outlined by Gleize et al. [46]. Briefly carotenoids, tocopherols and retinol were isolated from biological fluids by double hexane extraction with echinenone as the internal standard. Then, the extract was evaporated to dryness under nitrogen and dissolved in 200µL (for plasma) or 100µL (for FF) of a mixture of methanol and dichloromethane (65:35, v:v). A volume of 100 µL (for plasma) or 80 µL (for FF) was used for high-performance liquid chromatography (HPLC) analysis. Eleven fat-soluble micronutrients, including vitamin E (α- and γ-tocopherols), carotenoids subdivided into xanthophylls (β-cryptoxanthin, lutein, and zeaxanthin) and carotenes (phytoene, phytofluene, lycopene, α- and β-carotenes), along with vitamin A (retinol), were measured. The ThermoFisher Vanquish HPLC system comprised a VC-P20-A HPLC Pump, a VC-A12-A Automated Sample Injector, and a VF-D11-A-01 diode array detector (ThermoFisher Scientific, Waltham, Massachusetts, USA). Micronutrients were separated using a 250 × 4.6 mm, 5µm YMC C30 column preceded by a guard column (YMC Europe GMBH) and maintained at a constant temperature of 35°C. The mobile phase consisted of a gradient of methanol (A), methyl tert-butyl ether (B), and water (C). The run procedure of this mobile phase was at a flow rate of 1 mL/min with a variation of solvent gradient (A: B: C) as follows: 96: 2: 2 changing linearly to 18: 80: 2 in 27 min then returning to 96: 2: 2 from 31 to 35 min.

Phytoene, phytofluene, lycopene (all-trans plus cis isomers), other carotenoids, tocopherols and retinol were detected at 286, 348, 472, 450, 290 and 325 nm, respectively. Each molecule was identified by its retention time compared with pure standards and quantified using Chromeleon software (version 7.2.10), comparing peak areas with standard reference curves. Six-point external standard curves (ranging from 1 to 100 ng) were constructed for each compound, and micronutrient concentrations (nmol/L) were calculated using a linear regression equation.

Since plasma and FF compound concentrations are likely to vary over time due to diet, use of dietary supplements and changes in lifestyle, blastocyst transfers occurring more than six months after oocyte puncture were excluded to limit potential bias related to these temporal changes.

### Statistical analysis

Concentrations of carotenoids, vitamins E and A were described in terms of medians, interquartile ranges (IQR) and coefficients of variation (CV), and compared between plasma and FF using non-parametric paired Wilcoxon signed-rank tests, after rejection of distribution normality by Shapiro-Wilk tests. Relationships between compound concentrations within plasma, and between plasma and FF were assessed using Spearman’s correlation coefficients. We also calculated FF to plasma concentration ratios and assessed the correlation between the octanol–water coefficient (logP) and median FF to plasma ratios. LogP values for each molecule (computed by XLogP3 3.0) were retrieved from PubChem. Associations between compound concentrations and fruit and vegetable intakes were assessed using linear trend tests across increasing categories of consumption frequency. Comparisons of compound concentrations according to binary reproductive outcomes (success vs. failure) were performed using Wilcoxon rank-sum tests.

Associations between compound concentrations and IVF outcomes were assessed using different regression models according to the outcome variable type: linear regressions for continuous outcomes (LH, FSH and progesterone concentrations before oocyte puncture), binomial regressions for proportion outcomes (mature oocyte, fertilization, blastocyst, and embryo/blastocyst quality rates), and unconditional logistic regressions for binary outcomes (biochemical pregnancy at days 7, 9, and 14 PET, clinical pregnancy, and live birth). Associations with oocyte count outcomes (total number of oocytes and number of mature oocytes retrieved per patient) were further explored using multivariable negative binomial regression models. For linear regression models, log-transformations were applied to outcome variables to improve normality. All regression models were adjusted for age at oocyte puncture (<30, 30 - 34, ≥35 years) and BMI (<25, ≥25 kg/m²). Other factors were examined, including smoking status, ovarian stimulation protocol, type of treatment for pituitary suppression, etiology of infertility, and female reproductive pathologies, but were not retained as potential confounders in the final models. To facilitate comparisons across molecules and fluids, compound concentrations were rescaled using fixed increments defined separately for each compound in plasma and FF. Rounded increments close to half the IQR were chosen to capture meaningful variations within the observed distribution, while accounting for FF/plasma ratios to ensure consistency between systemic and intra-follicular compartments. Sensitivity analyses were performed after excluding outliers. Outliers were defined as values greater than the upper quartile plus 1.5 times the IQR. To focus on the most meaningful findings, only associations that remained statistically significant (two-sided p<5%) after sensitivity analyses were retained. All analyses were conducted using R software (version 4.2.3).

## RESULTS

### Baseline characteristics and dietary habits of the study population

Baseline characteristics of the 82 women are summarized in **Table 1**. The median age was 33.6 years, 48.8% of participants were overweight or obese (BMI ≥ 25 kg/m²) and 22.6% reported being smokers or having stopped less than one year prior inclusion. The main infertility etiologies were evenly distributed between female and male factors (35.4% each). In this population, 19.5% of infertility cases were classified as idiopathic. Among female pathologies, diminished ovarian reserve was the most frequent (22.0%), followed by endometriosis (6.1%). Most women (67.1%) lived in a village or in a dispersed housing rather than in a city center or a suburban district (32.9%); 24.3% reported living near an agricultural or industrial area. In terms of occupation, 53.7% were employees and 30.5% held executive or senior intellectual positions.

**Table 1.** Descriptive characteristics and socio-demographic data of the study population, FERTENOX study, 2021-2022 (n=82 women)

| Characteristics | N or Median | % or [IQR] |
| --- | --- | --- |
| Age at oocyte puncture (years), median [IQR] | 33.6 | [30.1; 38.0] |
| Age at oocyte puncture (years) |  |  |
| 23.6 to < 30 | 20 | 24.4 |
| $\geq 30$ to <35 | 27 | 32.9 |
| $\geq 35$ to 42.7 | 35 | 42.7 |
| Height (cm), median [IQR] | 163 | [160; 168] |
| Weight (kg), median [IQR] | 65 | [56; 76] |
| BMI* ($\text{kg/m}^2$), median [IQR] | 24.9 | [21.8; 28.4] |
| BMI ($\text{kg/m}^2$) | | |
| Underweight (17.7 to <18.5) | 4 | 4.9 |
| Normal weight ($\geq 18.5$ to <25) | 38 | 46.3 |
| Overweight ($\geq 25$ to <30) | 25 | 30.5 |
| Obesity ($\geq 30$ to <40) | 14 | 17.1 |
| Severe obesity ($\geq 40$ to 40.6) | 1 | 1.2 |
| Smoker status† |  |  |
| Current smokers (daily or occasional use) | 13 | 16.3 |
| Recent quitters (stopped $\leq 1$ year ago) | 5 | 6.3 |
| Former smoker (stopped >1 year ago) | 18 | 22.5 |
| Never smoker | 44 | 55.0 |
| **Etiology of infertility** |  |  |
| Female factor | 29 | 35.4 |
| Male factor | 29 | 35.4 |
| Female factor and male factor combined | 8 | 9.7 |
| Idiopathic | 16 | 19.5 |
| **Type of female infertility** |  |  |
| Diminished ovarian reserve | 18 | 22.0 |
| Tubal pathology | 4 | 4.9 |
| Diminished ovarian reserve + Tubal pathology | 1 | 1.2 |
| Endometriosis | 5 | 6.1 |
| PCOS | 4 | 4.9 |
| Dysovulation | 2 | 2.4 |
| PCOS + Dysovulation | 3 | 3.7 |
| **Current residence** |  |  |
| Village or dispersed housing | 55 | 67.1 |
| Suburban district (large city) | 15 | 18.3 |
| City center (large city) | 12 | 14.6 |
| **Proximity of residence, close to:** |  |  |
| An agricultural zone | 16 | 19.5 |
| An industrial zone | 2 | 2.4 |
| An industrial zone and an agricultural zone | 1 | 1.2 |
| A wastewater treatment plant and an agricultural zone | 1 | 1.2 |
| No reported proximity to any of the above | 62 | 75.7 |
| **Occupation<sup>†</sup>** |  |  |
| Craftswoman, shopkeeper, business owner | 5 | 6.1 |
| Executive or senior intellectual profession | 25 | 30.5 |
| Employee | 44 | 53.7 |
| Housewife | 3 | 3.7 |
| Unemployed | 5 | 6.1 |

*BMI, body mass index; IQR, interquartile range [25th percentile; 75th percentile]; PCOS, polycystic ovarian syndrome.*
*\*BMI was calculated as the ratio of reported weight and height from the questionnaire.*
*†Time since smoking cessation was missing in two women, so percentages are computed on a total of 80 participants.*
*\*In this study no woman was farmer operator or manual worker; and no woman were on disability or long-time illness.*

Dietary habits of patients are presented in **Table 2**. Most patients (64.6%) consumed vegetables at least once a day, including 43.9% who reported consuming them several times a day. In contrast, 56.0% consumed fruit less than once per day, and 20.7% reported consuming fruit several times a day. Almost half (47.6%) reported taking at least one vitamin or trace mineral supplement. Specifically, 20.7% reported taking vitamin E, 11.0% vitamin A (retinol), and 4.9% carotenoids (α/β-carotenes or astaxanthin). Additionally, several participants reported supplementing with other micronutrients having direct or indirect anti-or pro-oxidant activity, including zinc (29.3%), vitamin C (24.4%), selenium (20.7%), and copper (8.5%).

**Table 2.**
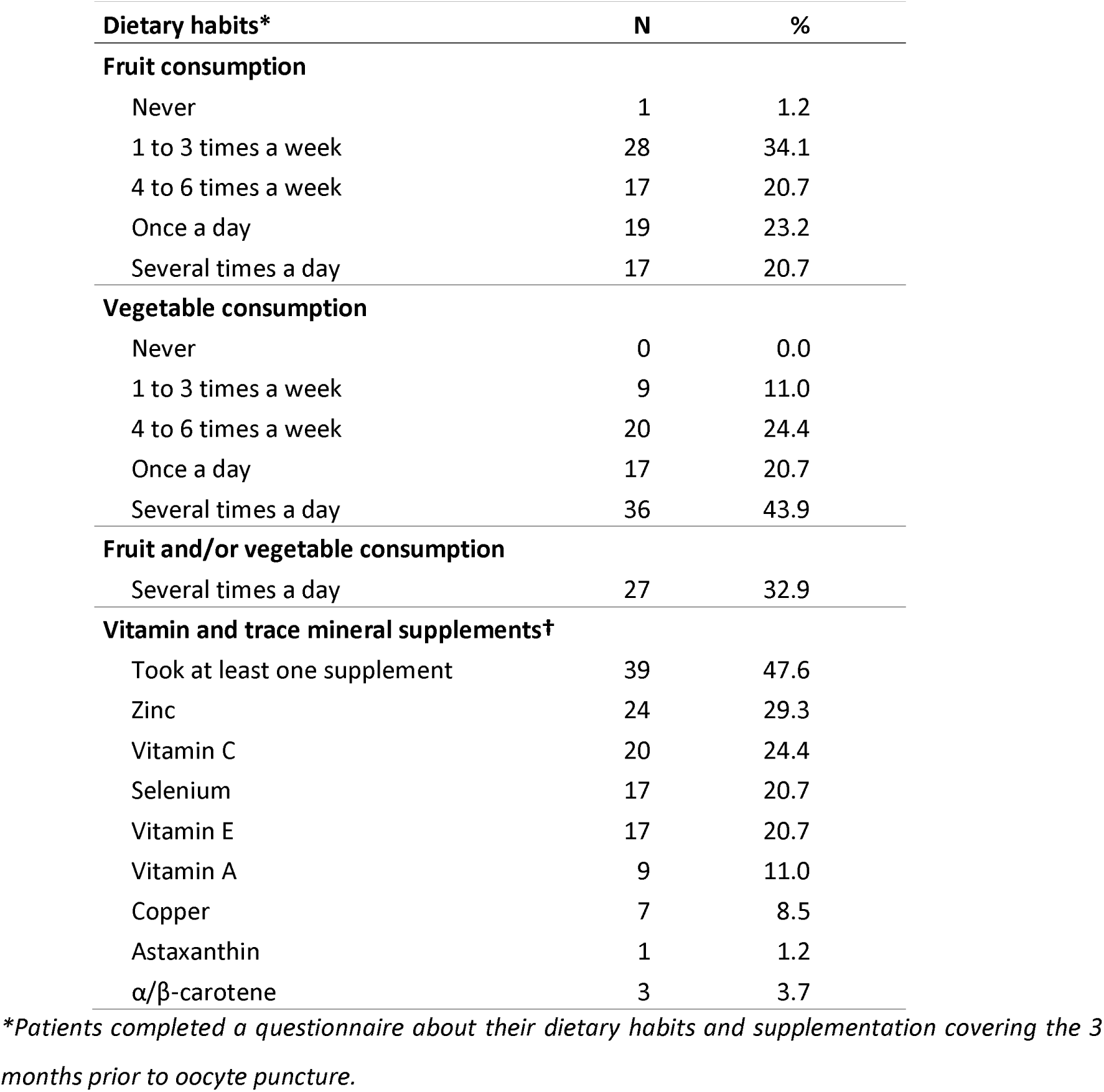
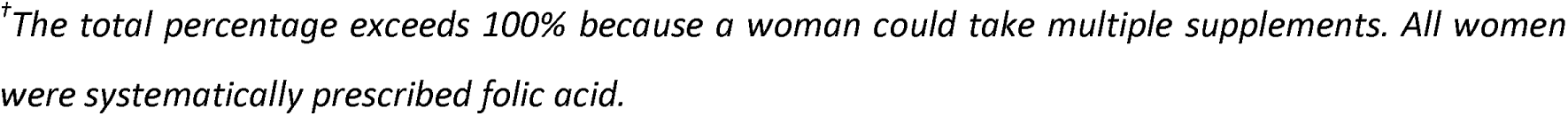
Dietary habits and supplementation of the study population, FERTENOX study, 2021-2022 (n=82 women)

| Dietary habits* | N | % |
| --- | --- | --- |
| **Fruit consumption** |  |  |
| Never | 1 | 1.2 |
| 1 to 3 times a week | 28 | 34.1 |
| 4 to 6 times a week | 17 | 20.7 |
| Once a day | 19 | 23.2 |
| Several times a day | 17 | 20.7 |
| **Vegetable consumption** |  |  |
| Never | 0 | 0.0 |
| 1 to 3 times a week | 9 | 11.0 |
| 4 to 6 times a week | 20 | 24.4 |
| Once a day | 17 | 20.7 |
| Several times a day | 36 | 43.9 |
| **Fruit and/or vegetable consumption** |  |  |
| Several times a day | 27 | 32.9 |
| **Vitamin and trace mineral supplements†** |  |  |
| Took at least one supplement | 39 | 47.6 |
| Zinc | 24 | 29.3 |
| Vitamin C | 20 | 24.4 |
| Selenium | 17 | 20.7 |
| Vitamin E | 17 | 20.7 |
| Vitamin A | 9 | 11.0 |
| Copper | 7 | 8.5 |
| Astaxanthin | 1 | 1.2 |
| $\alpha/\beta$-carotene | 3 | 3.7 |

\*Patients completed a questionnaire about their dietary habits and supplementation covering the 3 months prior to oocyte puncture.
<sup>†</sup>The total percentage exceeds 100% because a woman could take multiple supplements. All women were systematically prescribed folic acid.

### Carotenoid, vitamin E and A concentrations in plasma and follicular fluid

The concentrations of compounds in plasma and FF are presented in **Table 3**. Among the measured compounds, α-tocopherol was by far the most predominant in plasma, with a median concentration of 40,447 nmol/L, more than 20 times higher than that of retinol (1,869 nmol/L), the next most concentrated compound, followed by γ-tocopherol (772 nmol/L). The most abundant carotenoids in plasma were lutein (279.8 nmol/L), phytofluene (212.7 nmol/L), lycopene (197.3 nmol/L), and β-carotene (179.3 nmol/L). Phytoene plasma concentrations were markedly lower, with a median of 76.9 nmol/L. The highest CV in plasma was observed for β-cryptoxanthin (94.2%). Strong positive correlations between plasma compound concentrations were found for carotenoids (**Fig 1**): especially α- and β-carotenes (R = 0.79, p < 10^−5^), lutein and zeaxanthin (R = 0.72, p < 10^−5^), and phytoene and phytofluene (R = 0.71, p < 10^−5^).

**Figure 1.**
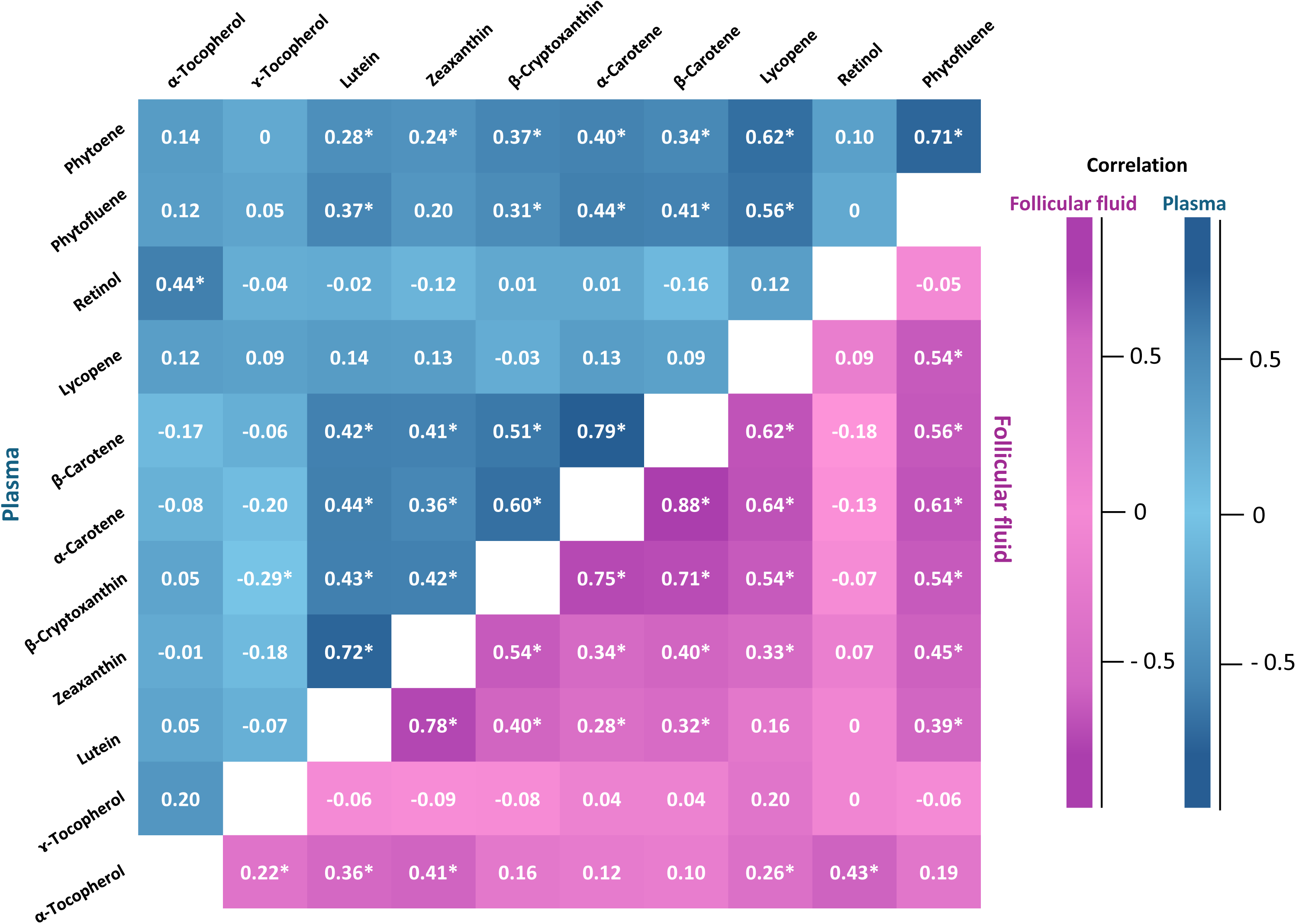
Spearman correlation matrix for carotenoids, vitamins E and A in plasma (blue) and in follicular fluid (pink), FERTENOX study, 2021-2022 (n=82 women) Compound concentrations were measured in plasma and follicular fluid samples after lipid extraction and using HPLC coupled to a photodiode array detector. * Spearman correlations with p-values less than 0.05.

**Table 3.**
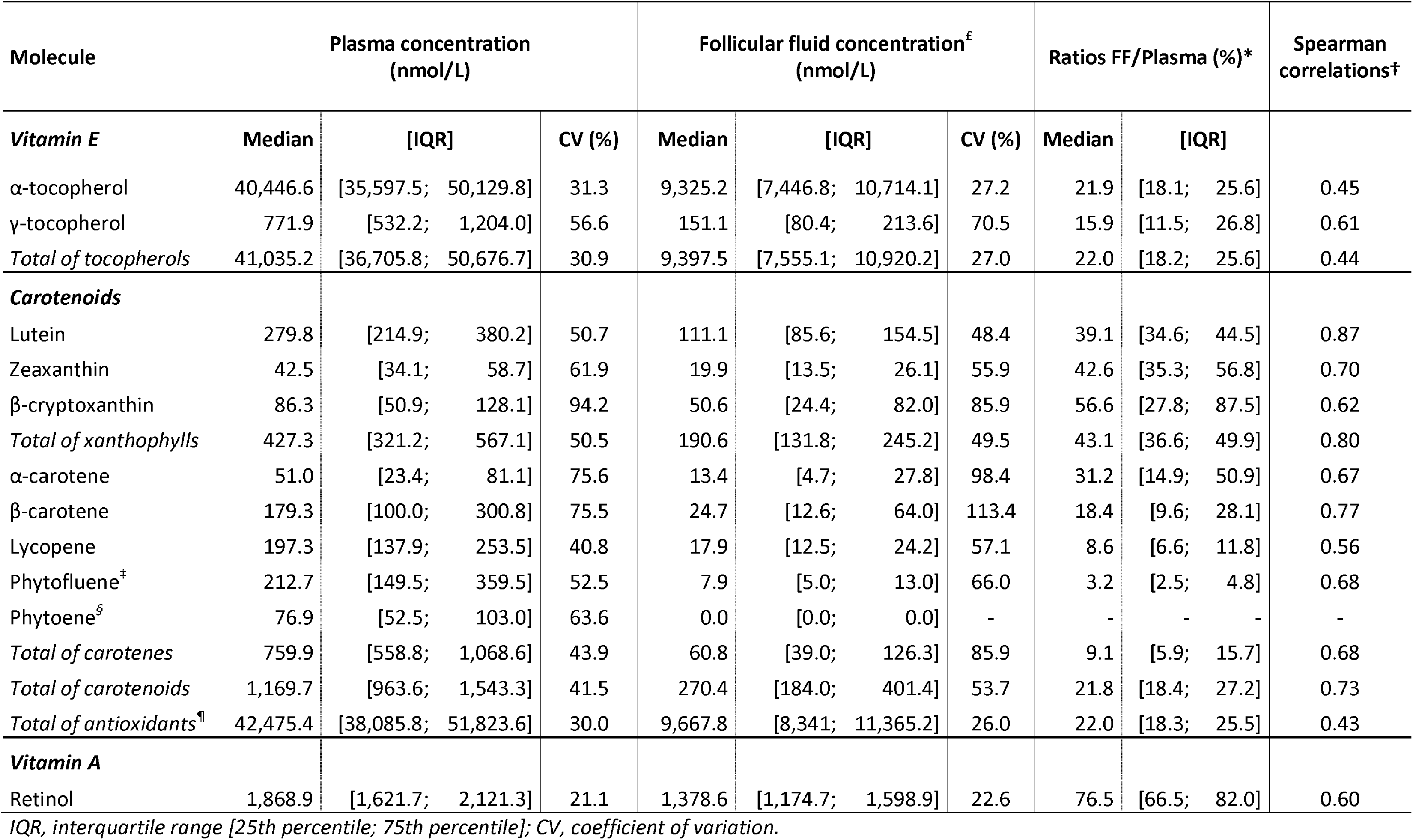
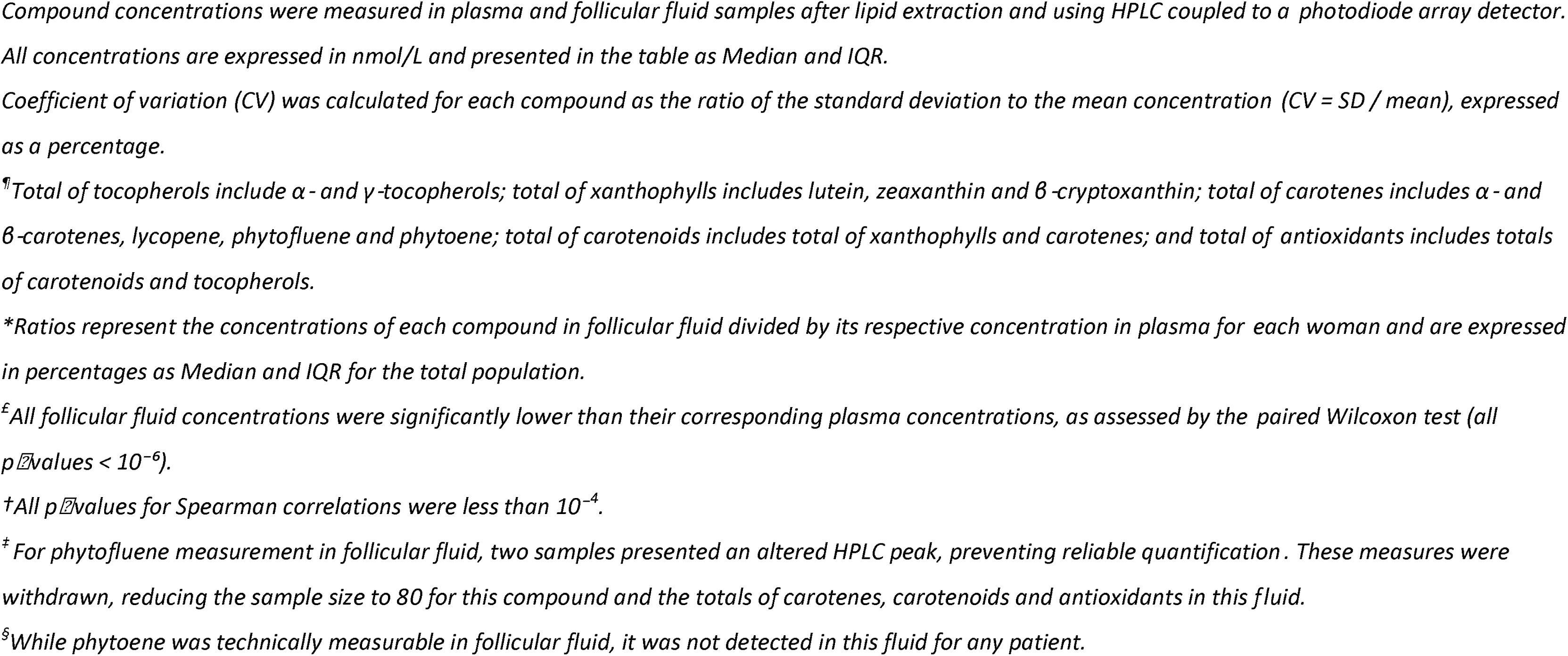
Carotenoids, vitamin E and vitamin A concentrations in plasma and follicular fluid of patients, FERTENOX study, 2021-2022 (n=82 women)

| Molecule | Plasma concentration (nmol/L) |  |  | Follicular fluid concentration<sup>£</sup> (nmol/L) |  |  | Ratios FF/Plasma (%)<sup>*</sup> |  | Spearman correlation<sup>†</sup> |
| --- | --- | --- | --- | --- | --- | --- | --- | --- | --- |
|  | Median | [IQR] | CV (%) | Median | [IQR] | CV (%) | Median | [IQR] |  |
| **Vitamin E** |  |  |  |  |  |  |  |  |  |
| α-tocopherol | 40,446.6 | [35,597.5; 50,129.8] | 31.3 | 9,325.2 | [7,446.8; 10,714.1] | 27.2 | 21.9 | [18.1; 25.6] | 0.45 |
| γ-tocopherol | 771.9 | [532.2; 1,204.0] | 56.6 | 151.1 | [80.4; 213.6] | 70.5 | 15.9 | [11.5; 26.8] | 0.61 |
| *Total of tocopherols* | 41,035.2 | [36,705.8; 50,676.7] | 30.9 | 9,397.5 | [7,555.1; 10,920.2] | 27.0 | 22.0 | [18.2; 25.6] | 0.44 |
| **Carotenoids** |  |  |  |  |  |  |  |  |  |
| Lutein | 279.8 | [214.9; 380.2] | 50.7 | 111.1 | [85.6; 154.5] | 48.4 | 39.1 | [34.6; 44.5] | 0.87 |
| Zeaxanthin | 42.5 | [34.1; 58.7] | 61.9 | 19.9 | [13.5; 26.1] | 55.9 | 42.6 | [35.3; 56.8] | 0.70 |
| β-cryptoxanthin | 86.3 | [50.9; 128.1] | 94.2 | 50.6 | [24.4; 82.0] | 85.9 | 56.6 | [27.8; 87.5] | 0.62 |
| *Total of xanthophylls* | 427.3 | [321.2; 567.1] | 50.5 | 190.6 | [131.8; 245.2] | 49.5 | 43.1 | [36.6; 49.9] | 0.80 |
| α-carotene | 51.0 | [23.4; 81.1] | 75.6 | 13.4 | [4.7; 27.8] | 98.4 | 31.2 | [14.9; 50.9] | 0.67 |
| β-carotene | 179.3 | [100.0; 300.8] | 75.5 | 24.7 | [12.6; 64.0] | 113.4 | 18.4 | [9.6; 28.1] | 0.77 |
| Lycopene | 197.3 | [137.9; 253.5] | 40.8 | 17.9 | [12.5; 24.2] | 57.1 | 8.6 | [6.6; 11.8] | 0.56 |
| Phytofluene<sup>‡</sup> | 212.7 | [149.5; 359.5] | 52.5 | 7.9 | [5.0; 13.0] | 66.0 | 3.2 | [2.5; 4.8] | 0.68 |
| Phytoene<sup>§</sup> | 76.9 | [52.5; 103.0] | 63.6 | 0.0 | [0.0; 0.0] | - | - | - | - |
| *Total of carotenes* | 759.9 | [558.8; 1,068.6] | 43.9 | 60.8 | [39.0; 126.3] | 85.9 | 9.1 | [5.9; 15.7] | 0.68 |
| *Total of carotenoids* | 1,169.7 | [963.6; 1,543.3] | 41.5 | 270.4 | [184.0; 401.4] | 53.7 | 21.8 | [18.4; 27.2] | 0.73 |
| *Total of antioxidants<sup>¶</sup>* | 42,475.4 | [38,085.8; 51,823.6] | 30.0 | 9,667.8 | [8,341; 11,365.2] | 26.0 | 22.0 | [18.3; 25.5] | 0.43 |
| **Vitamin A** |  |  |  |  |  |  |  |  |  |
| Retinol | 1,868.9 | [1,621.7; 2,121.3] | 21.1 | 1,378.6 | [1,174.7; 1,598.9] | 22.6 | 76.5 | [66.5; 82.0] | 0.60 |

<sup>£</sup>IQR, interquartile range [25th percentile; 75th percentile]; CV, coefficient of variation.
Compound concentrations were measured in plasma and follicular fluid samples after lipid extraction and using HPLC coupled to a photodiode array detector.
All concentrations are expressed in nmol/L and presented in the table as Median and IQR.
Coefficient of variation (CV) was calculated for each compound as the ratio of the standard deviation to the mean concentration ( $CV = SD / \text{mean}$ ), expressed
as a percentage.
<sup>¶</sup>Total of tocopherols include $\alpha$ - and γ-tocopherols; total of xanthophylls includes lutein, zeaxanthin and β-cryptoxanthin; total of carotenes includes $\alpha$ - and
β-carotenes, lycopene, phytofluene and phytoene; total of carotenoids includes total of xanthophylls and carotenes; and total of antioxidants includes totals
of carotenoids and tocopherols.
\*Ratios represent the concentrations of each compound in follicular fluid divided by its respective concentration in plasma for each woman and are expressed
in percentages as Median and IQR for the total population.

FF concentrations were 1.7 to 66 times lower than those measured in plasma, depending on the compound (all p < 0.0001, **Table 3**). Alpha-tocopherol remained the dominant compound (9,325 nmol/L in median), followed by retinol (1,379 nmol/L). Among carotenoids, the most abundant were lutein (111.1 nmol/L) and β-cryptoxanthin (50.6 nmol/L). Despite its high plasma concentration, phytofluene was the least abundant carotenoid in FF (7.9 nmol/L), and phytoene was not detected at all. The highest CVs in FF were found for β-carotene (113.4%) and α-carotene (98.4%). Correlations between FF concentrations were strongest between carotenoids (**Fig 1**): α- and β-Illcarotenes (R = 0.88, p < 10^−5^), lutein and zeaxanthin (R = 0.78, p < 10^−5^) and α-carotene and β-cryptoxanthin (R = 0.75, p < 10^−5^).

The highest FF/plasma ratio (**Table 3**) was observed for retinol (76.5%), followed by the xanthophylls β-cryptoxanthin (56.6%), zeaxanthin (42.6%), and lutein (39.1%). Carotenes had lower FF/plasma ratios, especially phytofluene (3.2%). Strong positive correlations between the two fluids (**Table 3**) were found, especially for lutein (R = 0.87, p < 0.0001), β-carotene (R = 0.77, p < 0.0001), total of carotenoids (R = 0.73, p < 10^−13^) and zeaxanthin (R = 0.70, p < 10^−13^). We observed an inverse relationship (R^2^ = 0.63) between partition coefficient (LogP) of the 11 compounds and their FF/plasma ratios **(S2 Fig)**. Retinol had the lowest logP value (5.7) but the highest FF/plasma ratio (76.5%), while lycopene and phytofluene had the highest logP values (15.6 and 15.4 respectively), but the lowest FF/plasma ratios (8.5 and 3.2 respectively). Although α- and γ-tocopherols had logP values (10.7 and 10.3 respectively) similar to those of xanthophylls such as zeaxanthin (10.9) and lutein (11), their FF/plasma ratios were half those of xanthophylls.

Furthermore, plasma concentrations of α-carotene, β-carotene and β-cryptoxanthin showed statistically significant positive linear trends with increasing consumption frequencies of both fruits (p<0.001) and vegetables (p≤0.001) (**S1 Table**). Plasma lutein and phytoene concentrations were also positively associated with more frequent fruit consumption (p≤0.01).

### Clinical characteristics, fertility outcomes and IVF success rates

Ovarian reserve, IVF treatment and hormonal characteristics of patients are detailed in **Table 4**. Most women (82.9%) had never undergone artificial insemination, while 17.0% had previously undergone at least three. Median hormone concentrations before oocyte puncture were 7.5 nmol/L for estradiol and 3.9 nmol/L for progesterone. Most participants underwent classical IVF (61.0%), 37.8% underwent ICSI and only one woman had the combination of the two technics (1.2%).

**Table 4.** Ovarian reserve, IVF treatment and hormonal characteristics of patients, FERTENOX study, 2021-2022 (n=82 women)

| Variable | n or Median | % or [IQR] | N |
| --- | --- | --- | --- |
| **Ovarian reserve and previous inseminations** |  |  |  |
| **Number of previous artificial inseminations** |  |  | 82 |
| 0 | 68 | 82.9 |  |
| 3 | 6 | 7.3 |  |
| 4 | 5 | 6.1 |  |
| 5 | 1 | 1.2 |  |
| 6 | 2 | 2.4 |  |
| **Antral follicle count**, median [IQR] | 20 | [12; 30] | 81 |
| **Anti-Müllerian Hormone** (ng/mL), median [IQR] | 2.5 | [1.2; 4.5] | 82 |
| **Type of hormonal treatment and IVF** |  |  |  |
| **Pituitary desensitization and ovarian stimulation** |  |  | 82 |
| GnRH Agonist + FSH | 1 | 1.2 |  |
| GnRH Agonist + FSH + LH | 5 | 6.1 |  |
| GnRH Antagonist + FSH | 48 | 58.5 |  |
| GnRH Antagonist + FSH + LH | 28 | 34.1 |  |
| **Type of IVF** |  |  | 82 |
| Classic IVF | 50 | 61.0 |  |
| ICSI | 31 | 37.8 |  |
| Classic IVF + ICSI | 1 | 1.2 |  |
| **Endometrial thickness and hormonal concentrations 2d before oocyte puncture** |  |  |  |
| **Endometrial thickness** (mm), median [IQR] | 10.0 | [8.9; 11.5] | 82 |
| **LH** (UI/L), median [IQR] | 2.5 | [1.3; 5.1] | 68 |
| **Estradiol** (nmol/L), median [IQR] | 7.5 | [4.9; 9.2] | 80 |
| **Progesterone** (nmol/L), median [IQR] | 3.9 | [1.8; 13.0] | 80 |

*IQR, Interquartile Range [25th percentile; 75th percentile]; PCOS, Polycystic Ovarian Syndrome; IVF, In Vitro Fertilization; ICSI, Intracytoplasmic Sperm Injection; GnRH, Gonadotropin-Releasing Hormone; FSH, Follicle Stimulating Hormone; LH, Luteinizing Hormone; n, sample size for each category; N, total sample size.*

Fertility and IVF outcomes are detailed in **Table 5** while **Fig 2** shows the number of participants at each stage of the IVF process from collected oocytes to the type of embryo transfer, and live birth. All participants were punctured a median number of 10 oocytes, and all had at least one mature oocyte. Both the total and mature oocyte counts were positively associated with plasma estradiol concentrations before oocyte puncture (both p<0.001) **(S2 Table)**. The median mature oocyte rate in this study population was 83.3%, and the median fertilization rate was 71.4%. Among the 81 women who underwent IVF within a year after oocyte puncture, two did not obtain any zygote. The median topIllquality embryo ratio was 12.5%, and the median fragmentation rate was 38.8%. At day 5, 77 participants had at least one blastocyst produced, with a median blastocyst rate at 57.1% and a topIllquality blastocyst ratio at 11.1%. Among the 76 women who underwent an embryo transfer within a year, 67 had a fresh embryo transfer and 9 had a cryopreserved embryo transfer. Of them, 33 achieved a positive βIllhCG test at day 9 PET, 25 had a biochemical pregnancy, and 20 resulted in a live birth. The median duration of singleton pregnancies was 39.5 weeks, and the median birth weight was 3,550 g.

**Figure 2.**
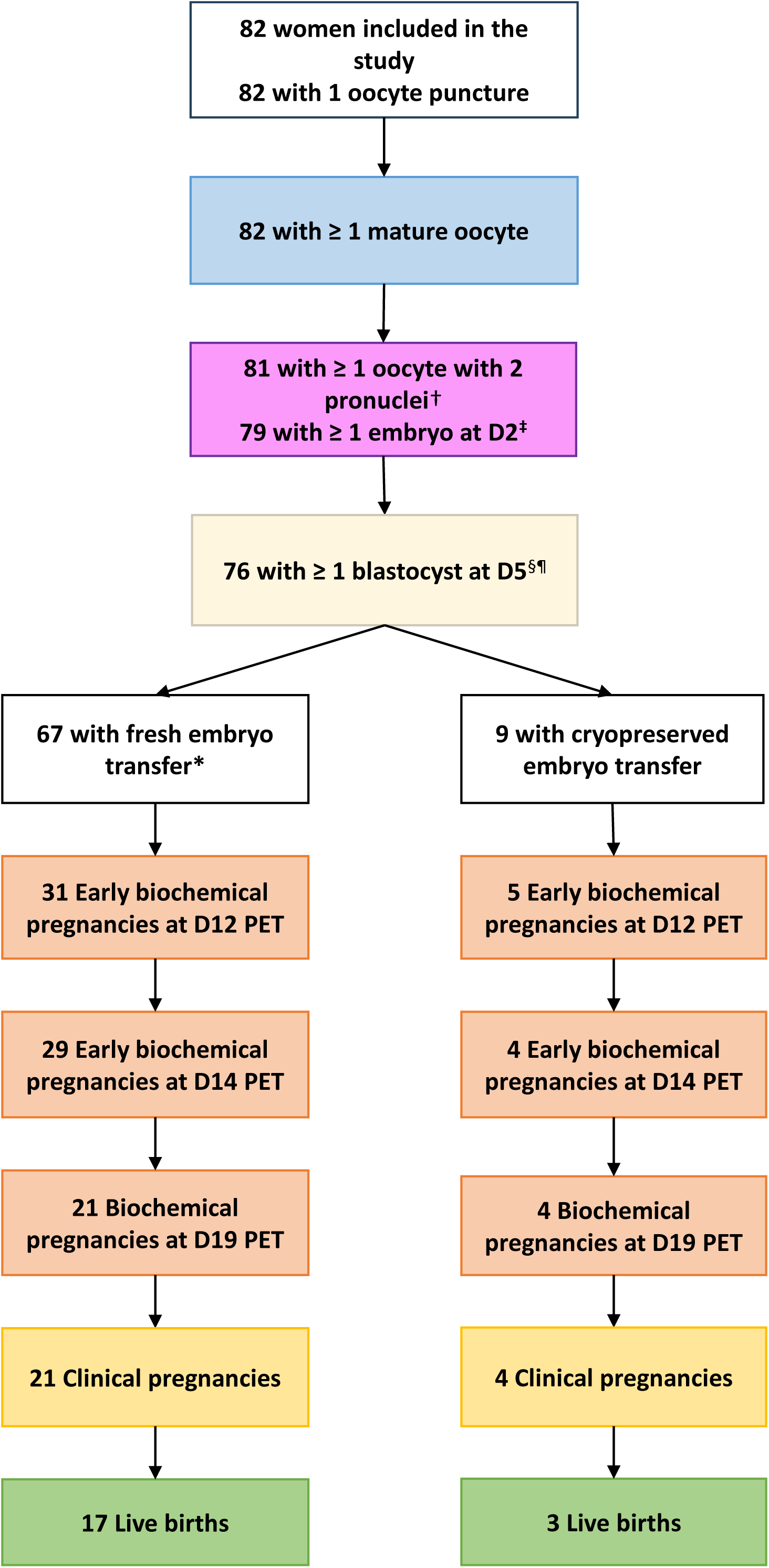
Flowchart of women according to in vitro fertilization outcomes, FERTENOX study 2021-2022 (n=82 women) D, day of oocyte puncture; PET, post embryo transfer; *Reasons for cryopreserved embryo transfer (n = 10): ovarian hyperstimulation (n = 7), hemoperitoneum (n = 1), Covid-19 positive test (n = 1), problem with catheter passage during transfer (n = 1). Fresh transfer: Embryo transfer performed 5 days after fertilization without freezing. Cryopreserved transfer: Embryo transfer conducted after the embryos have been frozen on day 5 after fertilization and later thawed for transfer. Early and biochemical pregnancies were determined based on beta-human Chorionic Gonadotropin (β-hCG) concentration in plasma, with early pregnancy defined as [β-hCG] ≥ 5 IU/L on day 7 post embryo transfer (D7 PET) and ≥ 10 IU/L on day 9 (D9 PET), and biochemical pregnancy confirmed for [β-hCG] > 1000 IU/L on day 14 (D14 PET). Clinical pregnancy was diagnosed at 8 weeks of amenorrhea with ultrasound when at least one fetus with a discernible heartbeat was detected. †One woman was excluded from the analysis at the fertilization stage because her IVF was performed more than one year after oocyte puncture. ^‡^Two patients did not have any embryo. ^§^Two patients did not have any blastocyst at day 5 after oocyte puncture.^¶^ One patient was excluded from the pregnancy outcomes analysis as her blastocyst transfer occurred more than one year after oocyte puncture.

**Table 5.**
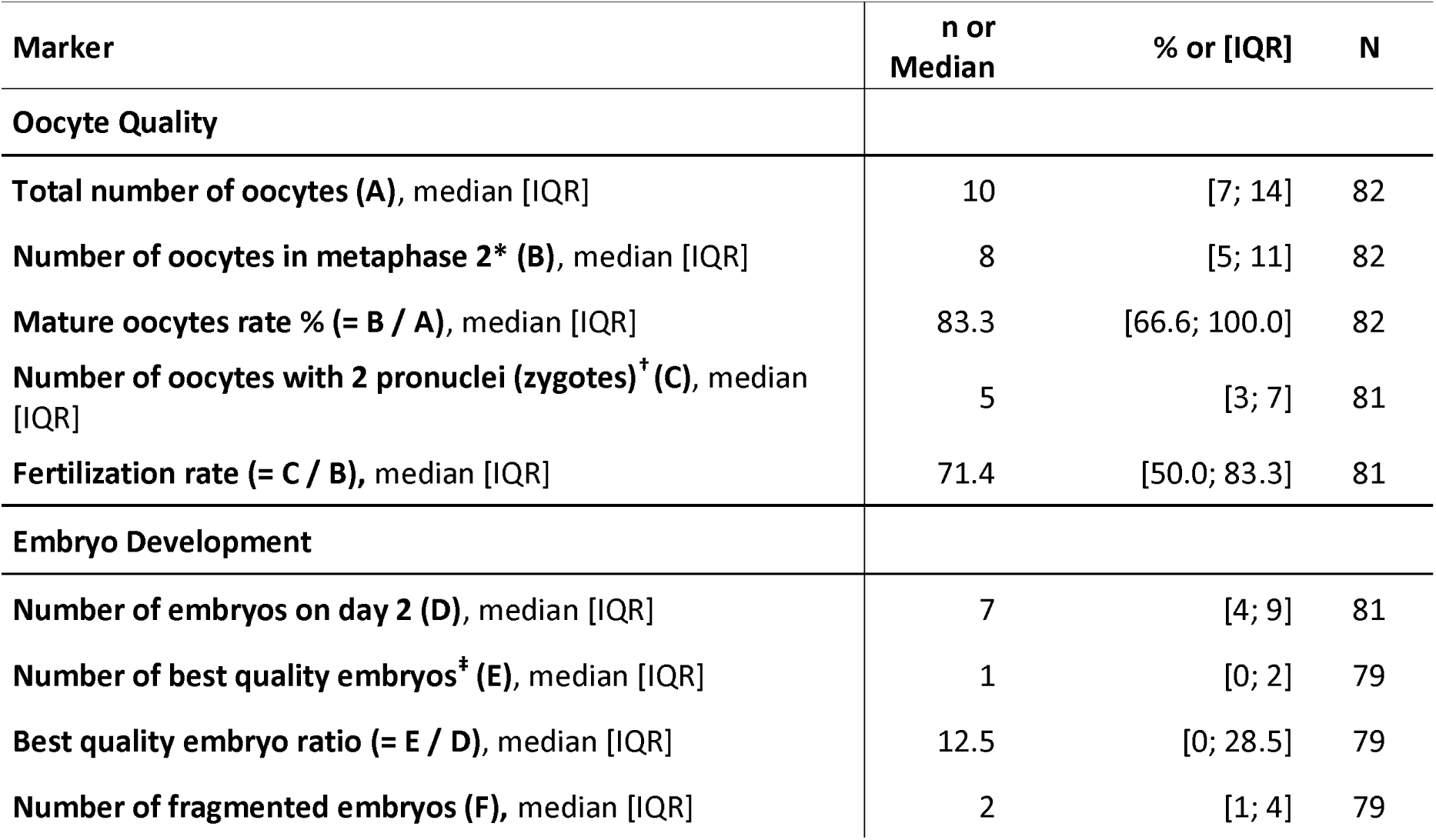
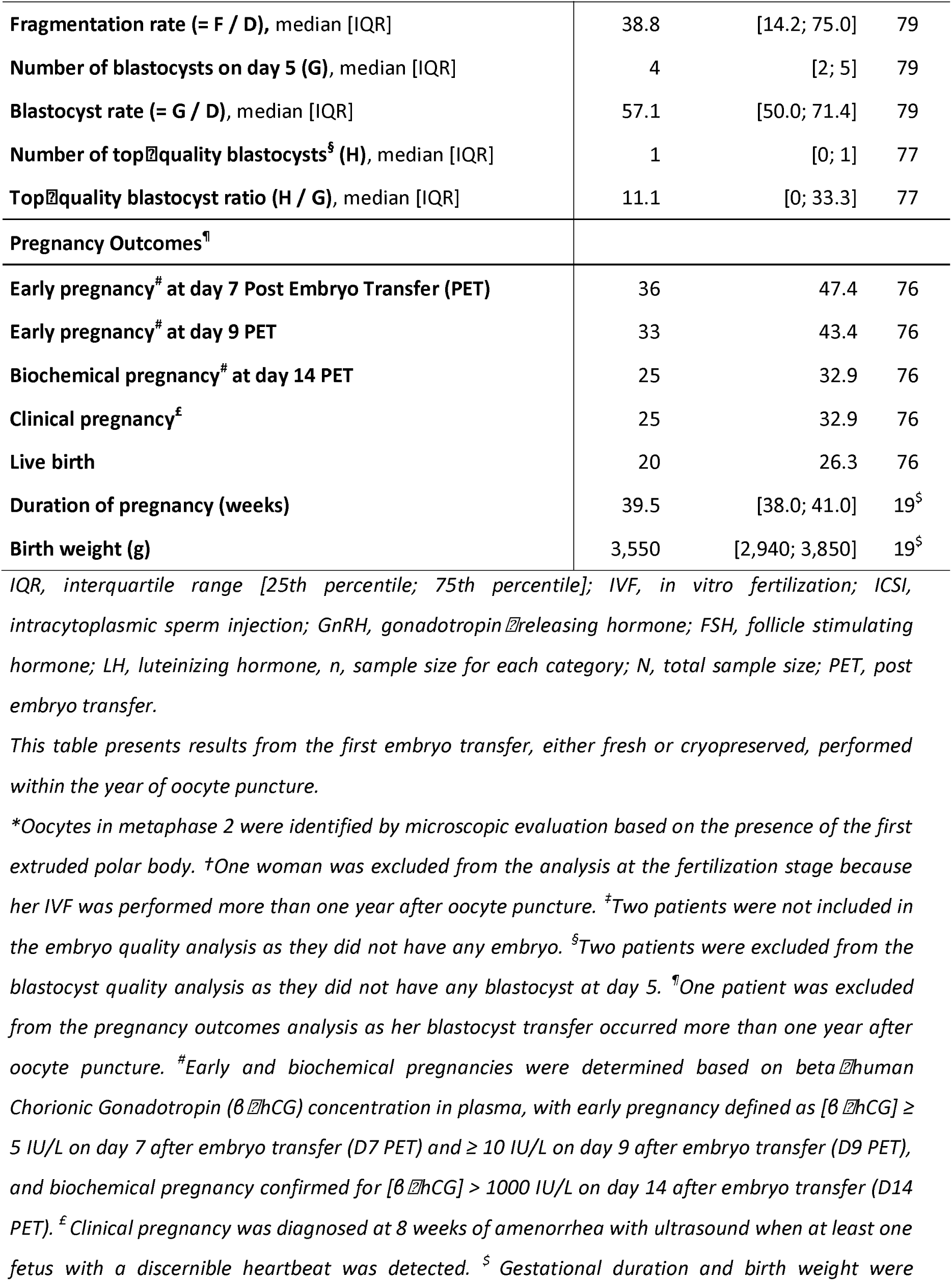
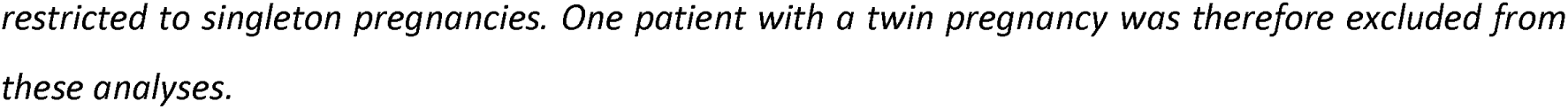
Oocyte quality, embryo development, and pregnancy outcomes of patients, FERTENOX study, 2021-2022 (n=82 women)

### Relationships between compound concentrations and IVF outcomes

Higher plasma concentrations of several compounds were observed in women without successful reproductive outcomes compared to women with successful outcomes (**S3 Table**). Statistically significant differences were found for early biochemical pregnancy at Day 7 PET (α-carotene and phytoene; both p≤0.03), clinical pregnancy (α-tocopherol, phytoene, total carotenoids, and total of antioxidants; all p≤0.03), and live birth (α-tocopherol, phytofluene, phytoene, and total of antioxidants; all p≤0.03).

**Table 6** presents adjusted results from the main analysis including outliers, highlighting only associations that remained statistically significant after exclusion of outliers in sensitivity analyses. All results, unadjusted and adjusted for age and BMI, exploring the relationships between compound levels in plasma or in FF and IVF outcomes are available without and after exclusion of outliers in **S4 and S5 Tables**, respectively. Several plasma compound concentrations showed statistically significant associations with hormonal, oocyte and clinical fertility outcomes, whereas the relationships observed with FF concentrations were fewer. In plasma, higher αIlltocopherol concentrations were positively associated with estradiol levels (p=0.008). Likewise higher β-cryptoxanthin levels were associated with increased progesterone concentrations (p=0.02). Among carotenoids, phytoene and phytofluene concentrations were positively associated with oocyte mature rate (p=0.01 and 0.03, respectively). After the fertilization stage, associations tended to reverse: plasma γIlltocopherol and phytofluene concentrations were inversely associated with fertilization rate (p<0.001) and topIllquality embryo rate (p=0.02), respectively, and γ-tocopherol and lycopene concentrations were positively associated with embryo fragmentation rate (p=0.01 and 0.03, respectively). At later stages, similar negative associations were observed between compound concentrations in plasma and pregnancy outcomes: higher αIllcarotene, αIlltocopherol and total antioxidant concentrations were associated with lower odds ratio of biochemical pregnancy at day 7 PET (p=0.02, 0.05 and 0.04, respectively) while elevated α-tocopherol, phytoene, and total antioxidant concentrations were also inversely related to clinical pregnancy (all p≤0.03) and live birth (all p≤0.04). There were no statistically significant associations between any xanthophyll concentration and any pregnancy outcome.

**Table 6.**
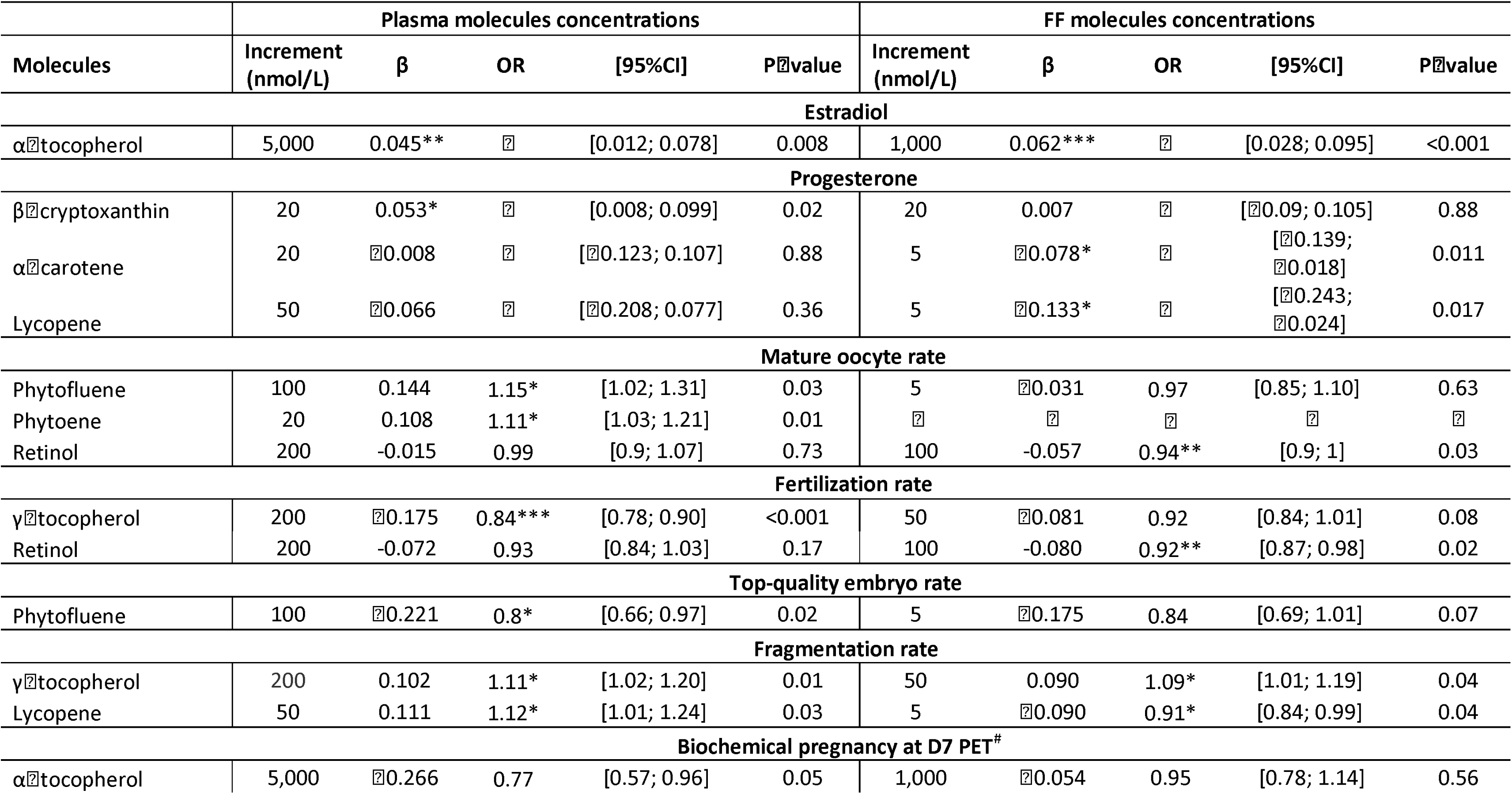
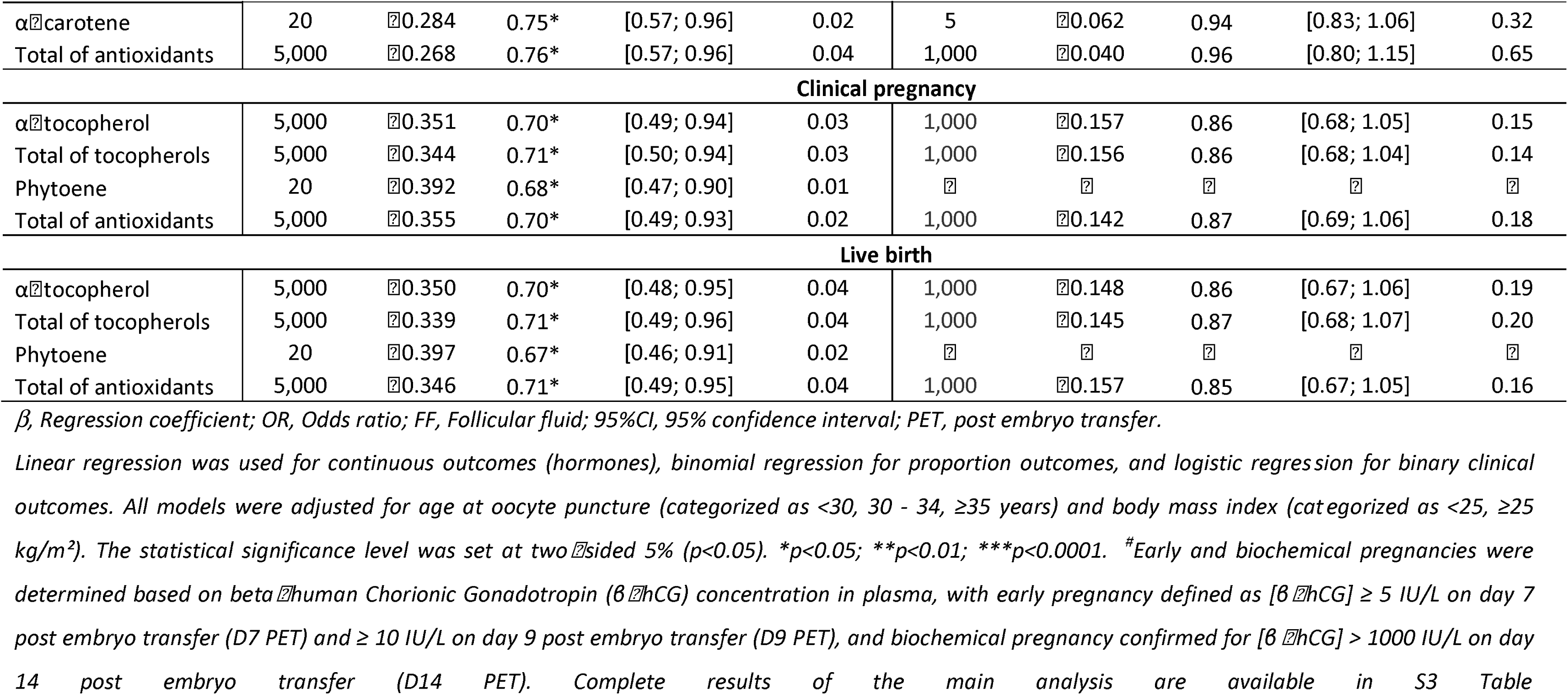
Selection of multivariable associations between carotenoid, vitamin E and vitamin A concentrations in plasma or follicular fluid and in vitro ilization outcomes, FERTENOX study, 2021-2022 (n=82 women)

For compound concentrations in FF, only few associations were statistically significant unlike in plasma (**Table 6**): FF α-tocopherol and γ-tocopherol concentrations were positively associated with estradiol levels (p<0.001) and embryo fragmentation rate (p=0.04), respectively. Conversely, FF retinol concentrations were inversely associated with oocyte maturation rate (p=0.03) and fertilization rate (p=0.02) but positively associated with the total number of oocytes retrieved per patient (p=0.012, **S2 Table**), while no statistically significant associations were observed between plasma retinol concentration and these outcomes. For lycopene concentrations in FF, we observed a negative association with fragmentation rate (p=0.04), unlike its plasma concentrations. For αIllcarotene and lycopene concentrations in FF, we found negative associations with progesterone levels (p=0.011 and 0.017, respectively), whereas these associations were not statistically significant in plasma. No other statistically significant associations were observed in FF.

## DISCUSSION

This study aimed to characterize the distribution of two tocopherols, eight carotenoids and retinol in plasma and FF from women undergoing IVF, and to explore their relationships with key markers of ovarian function, embryo development, and IVF and pregnancy success outcomes. All compounds were detected in both compartments, with lower concentrations in FF than in plasma. However, phytoene was not detected in FF, despite being present at higher concentrations in plasma than other carotenoids such as zeaxanthin and α-carotene, which were present in FF. Concentrations of α-tocopherol in both plasma and FF consistently showed positive relationships with estradiol level before oocyte puncture, while lycopene and γ-tocopherol exhibited inverse associations with embryo fragmentation rate that were observed in both biological compartments. Furthermore, other statistically significant associations were observed only between plasma concentrations of some compounds and outcomes: β-cryptoxanthin concentration in plasma was positively associated with plasma progesterone concentrations before oocyte puncture, phytofluene and phytoene concentrations were positively associated with mature oocyte rate, higher plasma levels of phytofluene and γ-tocopherol were associated with lower top-quality embryo rate and fertilization rate, respectively and α-tocopherol, phytoene, α-carotene and total antioxidant concentrations were inversely associated with several pregnancy success outcomes. Moreover, our study reported few statistically significant associations of IVF outcomes with FF compound concentrations only. All these relationships were negative and concerned lycopene and retinol. Finally, our results highlighted that statistically significant associations with markers of ovarian function, embryo development, and IVF or pregnancy success outcomes were mostly found with carotenes, in contrast to xanthophylls.

The most interesting finding regarding the relationships between compounds in biological fluids and female reproductive outcomes relied in the fact that beneficial associations were reported before the fertilization stage. Regarding hormonal concentrations measured before oocyte puncture, both plasma and FF concentrations of α-tocopherol were positively associated with estradiol levels, which is in line with a previous study in healthy premenopausal women [47]. In adult mouse, an increase in plasma estradiol was measured after a daily subcutaneous administration of α-tocopherol for six days [48]. Furthermore, in our study, plasma estradiol concentration before oocyte puncture was positively correlated with the number of retrieved oocytes and mature oocytes, in accordance with several previous studies in women who underwent IVF or ICSI [49–51]. Supplementation with α-tocopherol in the in vitro maturation medium did not enhance the maturation of ovine [52] or bovine [16] oocytes. However, a study demonstrated that α-tocopherol may exert a beneficial effect on the maturation of bovine oocytes only collected during summer, but not during spring [17]. In our study, α-tocopherol concentrations were not associated with the numbers of retrieved or mature oocytes, like a previous study in women [37] but contrary to another which found positive associations [40]. Further studies are needed to explore the role of α-tocopherol on ovarian response potentially mediated by estradiol.

Plasma phytofluene and phytoene, investigated here for the first time in relation to ART outcomes, were the only compound concentrations positively associated with the mature oocyte rate. These two carotenes are found in some fruits, vegetables and abundantly in tomatoes and tomato-enriched products. They are precursors of all the other carotenoids in nature [53] and act as effective antioxidants: with properties similar to those of α-tocopherol and β-carotene, but less than lycopene [44]. They can integrate into lipoproteins in vitro and reduced their oxidation, reduce oxidative damage in different cell types and modulate endogenous antioxidant defense via the Nuclear factor erythroid 2–related factor 2 / Antioxidant Response Element (Nrf2/ARE) pathway [54]. To our knowledge, there is currently no data on the role of phytoene and phytofluene in ovarian cells. Given their antioxidant activity, they might act similarly to other antioxidants by protecting oocytes and enhancing oocyte maturation by scavenging ROS, increasing antioxidant gene expression, preserving mitochondrial function or reducing lipid peroxidation in oocytes or cumulus-oocyte complexes (for review, [55,56]). In our study, the positive associations with mature oocyte rate were observed only with plasma compound concentrations and not with those in FF, especially since phytoene was not detected in FF. Given the presence of the follicular blood barrier, which may limit the transfer of certain carotenoids such as phytoene, these findings suggest that the observed relationships may reflect indirect systemic effects rather than a direct action within the follicular environment. Further investigations are required to decipher the potential roles and mechanisms of action of these carotenes on oocyte development.

Unexpectedly, statistically significant negative associations between several plasma compounds and success of fertilization, embryo development and pregnancy were reported in the present study. Indeed, plasma concentrations of γ-tocopherol, phytofluene, and lycopene were inversely associated with fertilization rate or top-quality embryo rate, and/or positively associated with embryo fragmentation rate, a deleterious indicator of embryo quality. In addition, higher plasma levels of α-tocopherol, α-carotene, phytoene and total antioxidants were associated with lower odds of biochemical pregnancy at D7 PET and/or clinical pregnancy and/or live birth.

While most studies reported no significant associations between plasma concentrations of these compounds and either fertilization success [38] or pregnancy status [39,40], Palini et al. [40] observed a positive association between β-carotene and pregnancy status. In our analysis, however, this carotenoid concentration was not statistically significantly associated with any embryo or pregnancy-related outcomes. Otherwise, our study found no statistically significant association between the FF concentrations of these compounds and embryo/blastocyst or pregnancy outcomes, except for γ-tocopherol, which showed a persistent positive association with the embryo fragmentation rate. These findings are consistent with previous studies which reported no statistically significant associations (all compounds [39], lutein/zeaxanthin and lycopene [40,42], α-tocopherol and lycopene [38] and β-carotene [42]). However, a few studies have reported positive associations with blastocyst count and embryo quality (α-tocopherol, [43]), or pregnancy status (α-tocopherol and β-carotene, [40]). Notably, only one study, consistently with our plasma data on γ-tocopherol, demonstrated a positive association between α-tocopherol (γ isomer not specified) and the embryo fragmentation rate [42]. These discrepancies across studies may stem from multiple sources of heterogeneity, including differences in exposure assessment or population characteristics or compound concentrations. While moderate antioxidant levels may support oocyte maturation and early embryo competence, excessive concentrations could disrupt the redox-sensitive signaling required for fertilization, cleavage, and implantation. Controlled levels of ROS are essential for physiological processes such as sperm–oocyte fusion, oocyte activation, and early embryogenesis [57]. An overly reduced environment, or an imbalance between maternal and paternal redox states, might therefore hinder optimal embryo development and implantation within the uterine milieu. Interestingly, Schweigert et al. [39] demonstrated that the ratios of FF to plasma concentrations of carotenoids and α-tocopherol were consistently higher in non-pregnant women compared to pregnant women, suggesting that increased transfer of these compounds into the FF may represent a detrimental factor in the context of IVF treatment. A comparative analysis of plasma and FF concentrations of these compounds from existing and future studies could provide valuable insights into their relationship with IVF outcomes. Additionally, these stage-specific differences may reflect the shift from an in vivo environment before fertilization to a standardized in vitro culture system after fertilization, which may attenuate inter-individual variability in micronutrient exposure, followed by a return to the in vivo maternal environment after embryo transfer. Finally, as carotenoids are established biomarkers of fruit and vegetable intake, and were positively associated with consumption frequencies in our study, an alternative explanation may involve exposure to dietary contaminants such as pesticides. Fruits and vegetables are a major source of pesticide exposure when conventionally grown, and evidence shows that consuming organic versions of these foods markedly reduces urinary pesticide residues [58]. However, for our specific population of women, data regarding the cultivation methods used for the fruits and vegetables (conventional or organic) were not available to substantiate this hypothesis and further investigations would be required.

With respect to retinol, no statistically significant associations were observed with its plasma concentration in the present study, as previously reported for fertilization success [38] and pregnancy status [39,40]. However, FF retinol concentrations in our study were positively associated with the total oocytes retrieved per patient, an indicator of ovarian stimulation response, while FF retinol concentrations were inversely associated with mature oocyte rate and fertilization rate, suggesting a potential dissociation of its role between ovarian response and oocyte competence and fertilization. Palini et al. [40] reported higher follicular retinol concentrations in women who did not achieve pregnancy, but other studies did not observe statistically significant associations with several IVF outcomes [38,39,42]. Furthermore vitamin A deficiency is known to be detrimental to maternal and fetal health, being associated with adverse outcomes such as preterm delivery and maternal anemia [59]. Overall, these heterogeneous findings highlight that the role of vitamin A in reproductive processes remains unclear and warrants further research, particularly in the context of IVF treatment.

Overall, the range of plasma concentrations of fat-soluble molecules in our study were comparable to those reported in some large European epidemiological cohorts [60–63], and in clinical studies of women undergoing IVF treatment [38–40]. Similarly, all FF compound concentrations were in the range of concentrations observed in previous studies [38–40,64–66] and significantly lower in FF than in plasma [39,40,65]. The reduced concentrations of compounds in FF compared with plasma are in line with the selective permeability of the blood–follicle barrier described by Browne et al. [67]. Similarly to Schweigert et al. [39] and Palini et al. [40], we observed higher FF/plasma ratios for xanthophylls compared with carotenes. This pattern could be partly explained by the selective transfer of micronutrients across the blood–follicle barrier: only molecules associated with high-density lipoproteins, such as α-tocopherol and more polar carotenoids (xanthophylls: lutein, zeaxanthin, β-cryptoxanthin), are efficiently transferred to the FF, whereas less polar carotenoids (α-carotene, β-carotene, lycopene, phytoene, phytofluene) carried primarily by low-density lipoproteins are less available [39,67]. Their physicochemical properties play an important role in their type-dependent lipoprotein distribution and transport and thus in this FF distribution. Indeed, we observed an inverse relationship between molecular hydrophobicity (LogP) and FF/plasma ratios, with more hydrophobic compounds showing lower transfer into the FF. For example, the observation that phytoene was undetectable in FF in our study is consistent with its low abundance in plasma compared to phytofluene or lycopene, which is partly explained by the fact that phytoene is an early biosynthetic precursor of other carotenoids [53], and its highly apolar chemical structure.

This study has several notable strengths. First, its sample size, although modest, is relatively large compared with most previous biomarker-based studies investigating fat-soluble molecules in reproductive settings (n=82 vs 77 [39], 50 [38], 25 [40], and 16 [65]). Second, the simultaneous assessment of these compounds in both plasma and FF provides complementary insights into systemic availability and the ovarian microenvironment. Importantly, this is, to our knowledge, the first study to explore phytoene and phytofluene concentrations in FF, thereby broadening the spectrum of carotenoids considered in reproductive biology. Another strength lies in the availability of dietary and lifestyle data obtained through standardized questionnaires. Although these data do not allow precise quantification of nutrient intake, they provide useful contextual information to explore potential determinants of micronutrient status, such as frequency of dietary intake and supplementation, smoking, or physical exercise and help to contextualize inter-individual variability. In addition, elements of study validation support the biological coherence of our findings: for instance, the frequency of fruit and vegetable consumption in our cohort was strongly correlated with plasma concentrations of specific carotenoids, consistent with previous studies showing that dietary intake of fruits and vegetables is directly associated with circulating carotenoid levels [68]. Finally, the large panel of IVF outcomes assessed provides a more comprehensive evaluation of the potential links between fat-soluble micronutrient status and reproductive processes than most previous studies.

Nevertheless, several limitations should be acknowledged. Although our cohort is one of the largest of its kind, the study population of 82 women remains relatively small, limiting statistical power, especially for specific subgroup analyses, e.g., by infertility subtypes. In addition, micronutrient concentrations were assessed at a single time point (oocyte puncture), which may not capture the fluctuations occurring throughout the ovarian stimulation cycle, as already described in women who have undergone a treatment for IVF/ICSI protocol [40] or even during phases of the menstrual cycle in healthy premenopausal women [47]. Male factors were not addressed in the present analysis although they also play a fundamental role in fertilization and embryo development. Another limitation concerns female pathologies: a few women in our cohort had underlying conditions such as endometriosis or PCOS, which are both characterized by elevated levels of ROS and oxidative stress in the follicular environment [57,66]. However, in our study, we did not observe statistically significant differences in compound concentrations or IVF outcomes according to these conditions, suggesting that their impact may be limited in this dataset. Furthermore, although some participants reported using other antioxidant supplements such as vitamin C, selenium, and zinc, this subgroup was too small to allow reliable statistical analyses, and we did not measure these compounds in biological fluids, although their activities could contribute to the associations observed with IVF outcomes.

## CONCLUSIONS

Our study provides a comprehensive characterization of fat-soluble micronutrients including the carotenoids phytoene and phytofluene in plasma and FF during IVF, highlighting that while α-tocopherol and some carotenes may support steroidogenesis or oocyte maturation, higher antioxidant concentrations do not consistently translate into improved fertilization, embryo development, or pregnancy outcomes. These findings underscore the complexity of redox regulation in reproduction. Importantly, our results do not support the assumption that increasing dietary carotenoid, vitamins E or A intakes or supplementation would necessarily improve IVF outcomes. Larger, well-designed studies are needed before any evidence-based recommendations regarding carotenoid or tocopherol intake can be established for women undergoing IVF. Future research should incorporate additional markers of oxidative stress, including enzymatic defenses such as superoxide dismutase, catalase, and glutathione peroxidase, alongside global indices like Total Antioxidant Capacity, and integrate male oxidative status to better capture the multifactorial nature of reproductive success.

## Data Availability

The datasets analyzed during the current study are embargoed until publications of main results by the research team and so are not publicly available. However, data are available from the corresponding author upon reasonable request.

## ACKNOWLEDGEMENTS

The authors are grateful to all patients for their participation in the study. We thank the medical team and the IVF laboratory staff of CHRU of Tours for monitoring the patient’s care pathway and for collecting biological samples and the lifestyle questionnaire. The authors specifically acknowledge Claire Vignault who helped VM and FG in drafting all the documents required for the study’s approval request to the ethics committee, Dayana González Bolaños for her contribution in processing the questionnaire data and first analysis of IVF outcomes, Sophie Thomas-Thiébaut for her contribution in analyzing vitamin and trace mineral supplements.

## LIST OF ABBREVIATIONS

ART: assisted reproduction technology
BMI: body mass index
CHRU: Centre Hospitalier Régional Universitaire (Regional University Hospital Centre)
CI: confidence interval
CV: coefficient of variation
FF: follicular fluid;
FSH: follicle-stimulating hormone
GnRH: gonadotropin-releasing hormone
hCG: human chorionic gonadotropin
HPLC: high-performance liquid chromatography
ICSI: intracytoplasmic sperm injection
IQR: interquartile range
IVF: in vitro fertilization
LH: luteinizing hormone
PCOS: polycystic ovary syndrome
PET: post embryo transfer
ROS: reactive oxygen species.

## Authors’ contributions

VM and FG conceptualized the epidemiological study and SE helped with this conceptualization. VM acquired the funding for the study. FG collected clinical data on patients. PB and CSH performed the quantification of micronutrient concentrations in biological fluids and helped in the analysis of micronutrient data in the two biological fluids. VM and ACMT performed data curation. AN, ACMT and VM performed data analysis and interpretation, and were major contributors in writing the manuscript. All authors participated in writing the manuscript and approved the final version.

## SUPPORTING INFORMATION

### Supplementary figures

**S1 Figure (.pdf): Timeline of FERTENOX study protocol (2021-2022)** This figure presents the standard steps of the In Vitro Fertilization (IVF) protocol (blue arrows) and the steps specific to the FERTENOX protocol. D, Day of oocyte puncture (= Day 0); d, day; TD, Transfer Day (corresponding to D+5d for fresh embryos transfers and later dates for cryopreserved embryo transfers); IVF, In Vitro Fertilization; β-hCG, beta-human Chorionic Gonadotropin; LH, Luteinizing Hormone. Early and biochemical pregnancies were determined based on β-hCG concentration in plasma, with early pregnancy defined as [β-hCG] ≥ 5 IU/L on day 7 post embryo transfer (PET) (D7 PET) and ≥ 10 IU/L on day 9 post embryo transfer (D9 PET), and biochemical pregnancy confirmed as [β-hCG] > 1000 IU/L on day 14 post embryo transfer (D14 PET).

**S2 Figure (.pdf): Relationship between differential solubility of compounds (logP) and median follicular fluid to plasma concentration ratios of carotenoids, vitamins E and A, FERTENOX study, 2021-2022 (n=82 women)** FF, follicular fluid; logP, Octanol–water partition coefficient, defined as the logarithm of the ratio of a compound’s concentration in octanol to its concentration in water (logP = log(C_octanol/C_water)), used as an indicator of lipophilicity; FF/Plasma ratio (%): Median ratio of follicular fluid to plasma concentrations for each compound; R², coefficient of determination from the linear regression model, representing the proportion of variability in FF/plasma ratios explained by the partition coefficient (LogP). The solid line represents a linear regression fit between logP and FF/plasma ratios. LogP values were computational estimates generated using the XLogP3 3.0 algorithm obtained from PubChem (https://pubchem.ncbi.nlm.nih.gov; accessed on 2026/04/03). PubChem Compound Identifiers (CID): retinol (CID: 445354), α-tocopherol (CID: 14985), γ-tocopherol (CID: 92729), zeaxanthin (CID: 5280899), lutein (CID: 5281243), β-cryptoxanthin (CID: 5281235), β-carotene (CID: 5280489), α-carotene (CID: 6419725), phytoene (CID: 5280784), phytofluene (CID: 6436722), and lycopene (CID: 446925).

### Supplementary tables

**S1 Table (.xlsx): Median plasma concentrations of carotenoids, vitamins E and A according to categories of fruit and vegetable consumption frequency, FERTENOX study, 2021–2022 (n=82 women)** IQR, interquartile range [25th percentile; 75th percentile]; 95%CI, 95% confidence interval. **p<0.05; *p<0.10. ^£^Patients completed a questionnaire about their dietary habits covering the 3 months prior to oocyte puncture. ^†^P-values were obtained from linear regression models of micronutrient concentrations over categories of consumption frequencies of fruit and vegetables coded as 1 to 4 as follows: 1 = Less than once per week (reference category), 2 = 1–3 times per week, 3 = Once per day, and 4 = Several times per day. ^‡^β coefficients represent the change in log-transformed compound concentrations for each one-category increase in consumption frequency.

**S2 Table (.xlsx): Multivariable† associations between plasma and follicular fluid carotenoid, vitamin E and A concentrations and hormonal concentrations and the number of total and mature oocytes retrieved per patient, FERTENOX study, 2021–2022 (n=82 women)** β, regression coefficient representing the estimated change in the log count of oocytes per specified increment of each compound concentration; IRR, incidence rate ratio, equal to exp(β) reflecting the multiplicative change in the expected number of oocytes for each increment in compound concentration; 95%CI, 95% confidence interval. **p<0.05; *p<0.10.

†All models are adjusted for age at oocyte puncture (categorized as <30, 30 – 34, ≥35 years) and BMI (categorized as <25, ≥25 kg/m²). ^#^Compound concentrations were rescaled using fixed increments (≈ half the interquartile range), to ensure comparability between compound and consistency across biological fluids. ^‡^Hormonal concentrations (estradiol and progesterone) were log-transformed and analyzed using multivariable linear regression models.

**S3 Table (.xlsx): Concentrations of carotenoids, vitamins E and A (Median [IQR]) in plasma and follicular fluid in women according to success or failure for early biochemical pregnancies, clinical pregnancy and live birth, FERTENOX study, 2021-2022 (n=76 women)** IQR, interquartile range [25th percentile; 75th percentile]; N, total sample size; PET, post embryo transfer.

*Early and biochemical pregnancies were determined based on beta-human Chorionic Gonadotropin (β-hCG) concentration in plasma, with early pregnancy defined as [β-hCG] ≥ 5 IU/L on day 7 post embryo transfer (D7 PET) and ≥ 10 IU/L on day 9 post embryo transfer (D9 PET), and biochemical pregnancy confirmed for [β-hCG] > 1000 IU/L on day 14 post embryo transfer (D14 PET). ^£^Clinical pregnancy was diagnosed at 8 weeks of amenorrhea with ultrasound when at least one fetus with a discernible heartbeat was detected. ^$^P-values were calculated using Wilcoxon rank-sum tests to compare compound concentrations between outcome groups (success vs. failure). †Relative difference (%) = (median_success − median_failure) / median_failure × 100.

**S4 Table (.xlsx): Unadjusted and adjusted† multivariable associations between carotenoid, vitamin E and vitamin A concentrations in plasma or in follicular fluid and in vitro fertilization outcomes, FERTENOX study, 2021-2022 (n=82 women)** LH, luteinizing hormone; β, regression coefficient; 95%CI, 95% confidence interval; OR, odds ratio; PET, post embryo transfer. **p<0.05; *p<0.10.

Linear regression models were used for continuous outcomes (LH, estradiol, and progesterone concentrations, log-transformed); binomial logistic regression models were used for clinical outcomes reported as rates (mature oocyte, fertilization, top-quality embryo, fragmentation, blastocyst and top-quality blastocyst rates); and logistic regression models were used for binary outcomes (biochemical and clinical pregnancies and live birth). ^†^All models were adjusted for age at oocyte puncture (categorized as <30, 30 - 34, ≥35 years) and BMI (categorized as <25, ≥25 kg/m²). ^#^Compound concentrations were rescaled using fixed increments (≈ half the interquartile range), to ensure comparability between compounds and consistency across biological fluids. ^$^Early and biochemical pregnancies were determined based on beta-human Chorionic Gonadotropin (β-hCG) concentration in plasma, with early pregnancy defined as [β-hCG] ≥ 5 IU/L on day 7 post embryo transfer (D7 PET) and ≥ 10 IU/L on day 9 post embryo transfer (D9 PET), and biochemical pregnancy confirmed for [β-hCG] > 1000 IU/L on day 14 post embryo transfer (D14 PET). ^¶^Clinical pregnancy was diagnosed at 8 weeks of amenorrhea with ultrasound when at least one fetus with a discernible heartbeat was detected. This outcome overlapped the outcome “Biochemical pregnancy at Day 14 PET”.

**S5 Table (.xlsx): Unadjusted and adjusted† multivariable associations between carotenoid, vitamin E and vitamin A concentrations in plasma or in follicular fluid and fertility outcomes after exclusion of outliers, FERTENOX study, 2021-2022 (n=82 women)** LH, luteinizing hormone; β, regression coefficient; 95%CI, 95% confidence interval; OR, odds ratio; PET, post embryo transfer. **p<0.05; *p<0.10.

Linear regression models were used for continuous outcomes (LH, estradiol, and progesterone concentrations, log-transformed); binomial logistic regression models were used for clinical outcomes reported as rates (mature oocyte, fertilization, top-quality embryo, fragmentation, blastocyst and top-quality blastocyst rates); and logistic regression models were used for binary outcomes (biochemical and clinical pregnancies and live birth). ^†^All models were adjusted for age at oocyte puncture (categorized as <30, 30 - 34, ≥35 years) and BMI (categorized as <25, ≥25 kg/m²).

^£^Outliers were defined as compound concentration values exceeding the upper quartile plus 1.5 time the interquartile range. No values were below the lower quartile minus 1.5 time the interquartile range. Excluded values were from 1 patient for α-tocopherol, 1 patient for zeaxanthin, 2 patients for β-cryptoxanthin, 1 patient for phytoene, and 2 patients for lutein in plasma; and 1 patient for zeaxanthin, 1 patient for β-cryptoxanthin, and 1 patient for α-carotene in follicular fluid. ^#^Compound concentrations were rescaled using fixed increments (≈ half the interquartile range), to ensure comparability between compounds and consistency across biological fluids.

^$^Early and biochemical pregnancies were determined based on beta-human Chorionic Gonadotropin (β-hCG) concentration in plasma, with early pregnancy defined as [β-hCG] ≥ 5 IU/L on day 7 post embryo transfer (D7 PET) and ≥ 10 IU/L on day 9 post embryo transfer (D9 PET), and biochemical pregnancy confirmed for [β-hCG] > 1000 IU/L on day 14 post embryo transfer (D14 PET). ^¶^Clinical pregnancy was diagnosed at 8 weeks of amenorrhea with ultrasound when at least one fetus with a discernible heartbeat was detected. This outcome overlapped the outcome “Biochemical pregnancy at Day 14 PET”.

## Notes

### Competing Interest Statement

The authors have declared no competing interest.

### Clinical Trial

NCT04866329

### Author Declarations

The ethics committee Comite de Protection des Personnes of Ile‑de‑France gave ethical approval for this work.

