## Supplementary material for "Plasma and follicular fluid concentrations of carotenoids, tocopherols and retinol in a French population of women undergoing in vitro fertilization: a monocentric non-interventional study": S1 Figure

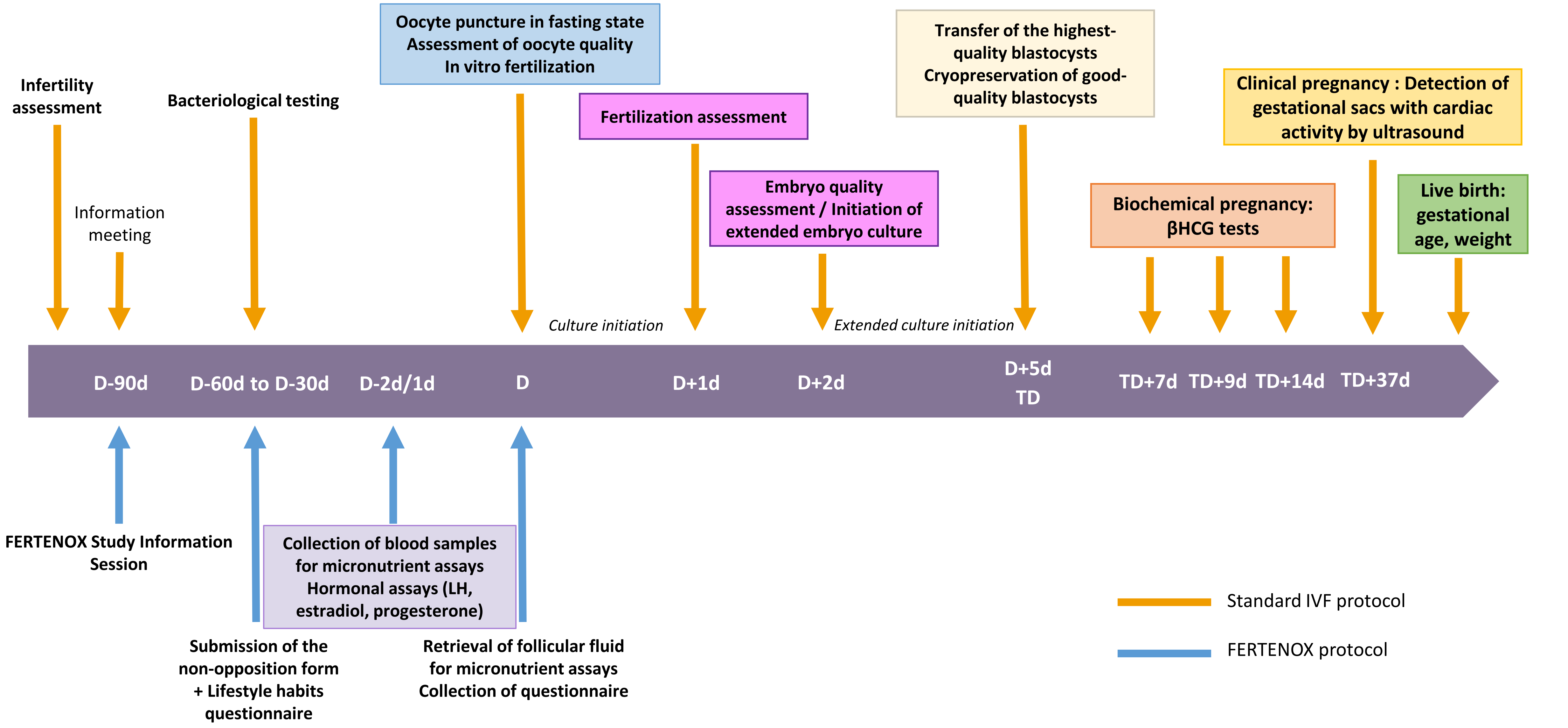


**S1 Figure: Timeline of FERTENOX study protocol (2021-2022)**

This figure presents the standard steps of the In Vitro Fertilization (IVF) protocol (blue arrows) and the steps specific to the FERTENOX protocol.

D, Day of oocyte puncture (= Day 0); d, day; TD, Transfer Day (corresponding to D+5d for fresh embryos transfers and later dates for cryopreserved embryo transfers); IVF, In Vitro Fertilization; β-hCG, beta-human Chorionic Gonadotropin; LH, Luteinizing Hormone.

Early and biochemical pregnancies were determined based on β-hCG concentration in plasma, with early pregnancy defined as [β-hCG] ≥ 5 IU/L on day 7 post embryo transfer (PET) (D7 PET) and ≥ 10 IU/L on day 9 post embryo transfer (D9 PET), and biochemical pregnancy confirmed as [β-hCG] > 1000 IU/L on day 14 post embryo transfer (D14 PET).
