## Supplementary material for "Plasma and follicular fluid concentrations of carotenoids, tocopherols and retinol in a French population of women undergoing in vitro fertilization: a monocentric non-interventional study": S2 Figure

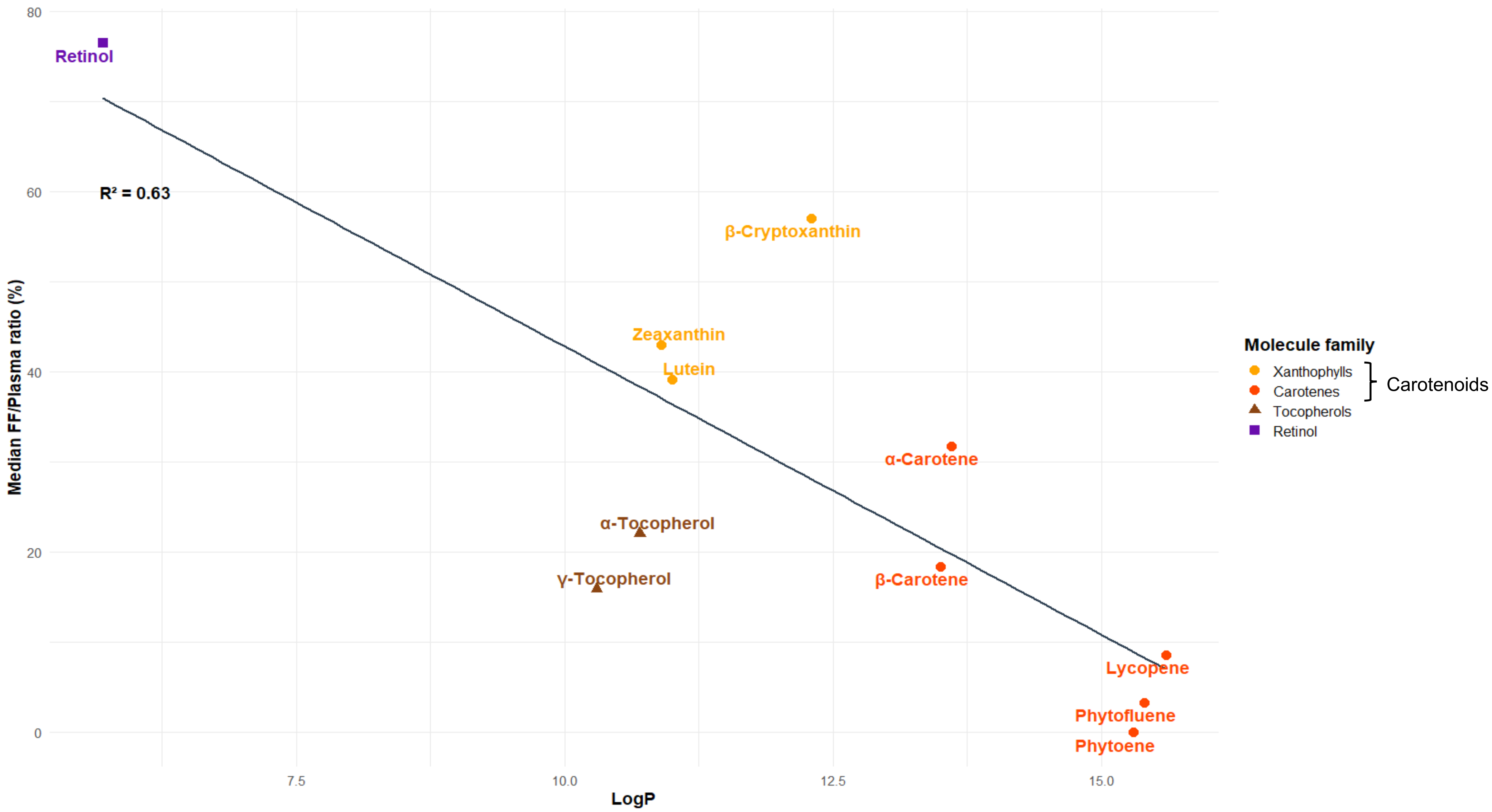


**Supplementary Figure 2. Relationship between differential solubility of compounds (logP) and median follicular fluid to plasma concentration ratios of carotenoids, vitamins E and A, FERTENOX study, 2021-2022 (n=82 women)**

FF, Follicular fluid; logP, Octanol–water partition coefficient, defined as the logarithm of the ratio of a compound's concentration in octanol to its concentration in water (logP = log(C octanol/C_water)), used as an indicator of lipophilicity; FF/Plasma ratio (%) :Median ratio of follicular fluid to plasma concentrations for each compound; $R^2$, coefficient of determination from the linear regression model, representing the proportion of variability in FF/plasma ratios explained by the partition coefficient (LogP). The solid line represents a linear regression fit between logP and FF/plasma ratios.

LogP values were computational estimates generated using the XLogP3 3.0 algorithm obtained from PubChem (https://pubchem.ncbi.nlm.nih.gov; accessed on 2026/04/03). PubChem Compound Identifiers (CID): retinol (CID: 445354), α-tocopherol (CID: 14985), γ-tocopherol (CID: 92729), zeaxanthin (CID: 5280899), lutein (CID: 5281243), β-cryptoxanthin (CID: 5281235), β-carotene (CID: 5280489), α-carotene (CID: 6419725), phytoene (CID: 5280784), phytofluene (CID: 6436722), and lycopene (CID: 446925).
